# The effect of highly effective modulator therapy on systemic inflammation in cystic fibrosis

**DOI:** 10.1101/2024.07.25.24310916

**Authors:** Rosemary E Maher, Urszula Cytlak-Chaudhuri, Saad Aleem, Peter J Barry, Daniel Brice, Eva Caamaño-Gutiérrez, Kimberley Driver, Edward Emmott, Alexander Rothwell, Emily Smith, Mark Travis, Dave Lee, Paul S McNamara, Ian Waller, Jaclyn A Smith, Andrew M Jones, Robert W Lord

## Abstract

**Background:** Despite significant clinical improvements, there is evidence of persisting airway inflammation in people with cystic fibrosis established on Elexacaftor/tezacaftor/ivacaftor (ETI) therapy. As CF is a multi-system disease, systemic immune profiles can reflect local inflammation within the lungs and other organs. Understanding systemic inflammation after ETI therapy may reveal important translational insights. This study aims to profile systemic inflammatory changes and relate these to the well-documented improvements observed with ETI therapy.

**Methods:** We conducted a single-centre longitudinal study with 57 CF subjects initiating ETI therapy. All participants were Phe508del homozygous or Phe508del/minimal function. Blood samples were collected pre-ETI and 3-12 months post-therapy initiation. Analyses included mass spectrometry-based proteomics, a multiplex immunoassay, and flow cytometry for peripheral immune cell counts and phenotype. Controls samples were provided by 29 age-matched healthy controls.

**Results:** Systemic inflammation reduced with ETI therapy; however, the immune profile remained distinct from healthy controls. ETI reduced neutrophil counts and was associated with a more mature, less inflammatory phenotype, as well as a shift toward an immune resolving state associated with increased CD206 expression. Cytokines known to influence neutrophil levels reduced with therapy. Despite ETI therapy, neutrophil and monocyte counts remained elevated compared to healthy controls. There was no obvious association between the ETI-related improvements in systemic inflammation and lung function.

**Conclusions:** Patients with CF show evidence of persisting systemic inflammation despite ETI therapy, this may have long term potentially adverse effects on respiratory and other organ systems.

## INTRODUCTION

Elexacaftor-Tezacaftor-Ivacaftor (ETI) therapy has led to dramatic improvements in cystic fibrosis (CF) lung disease across various measures, including lung function, radiology, and microbiology (1–4). However, ETI reduces but does not fully resolve airway inflammation (4, 5). This persistent airway inflammation may have implications for long-term lung health, raising concerns about the risks of progressive lung function decline and ongoing exacerbations, both of which could adversely impact quality of life and overall life expectancy (6, 7).

Systemic immune profiles may reflect airway inflammation, as inflammatory mediators released, and immune cells recruited to and from the airway are found within the peripheral circulation. Importantly, the systemic immune profile could also be influenced by inflammation within other organs affected by cystic fibrosis, such the gastro-intestinal and hepatobiliary systems (8). However, better understanding the relationship between systemic inflammation and CF lung disease would have obvious translational potential, aiding in the develop of blood-based surrogate markers of airway inflammation and novel precision systemic therapies. Furthermore, chronic systemic inflammation in other conditions has been associated with increased risk of cardiovascular disease, cancer and neurodegenerative disorders, that can adversely affect long-term outcome (9). There are concerns that these complications may become more prevalent in an aging CF population.

Systemic inflammation reduces with ETI, as evidenced by lower levels of circulating immune cells and inflammatory cytokines (10–14). However, it is not established whether, similar to the effect within the airway, systemic inflammation fully resolves with ETI therapy. This study aimed to provide a detailed description of the effect of ETI therapy on systemic inflammation and to establish how these changes relate to improvements seen in lung function. Employing a multimodal approach, we profiled systemic inflammation, spanning both cellular and molecular dimensions.

## MATERIALS AND METHODS

### Study design

This previously described prospective observational single-centre study recruited CF subjects from a large UK specialist CF centre (5). Ethical approval was granted to the Manchester Allergy, Respiratory and Thoracic Surgery (ManARTS) Biobank by the Northwest Haydock Research Ethics Committee (20/NW/0302). CF participants possessed two Phe508del alleles, or one Phe508del and a CFTR modulator unresponsive allele. Samples for analysis were collected from CF participants before (pre-ETI) and after ETI was commenced and ongoing at the time of follow-up (post-ETI). All participants provided written informed consent and were over 18 years, with a confirmed CF diagnosis based on genetic testing and/or sweat testing, along with typical phenotypic features, and deemed clinically stable by the medical team. Exclusion criteria included pregnancy and organ transplantation. The healthy control group comprised adult non-smokers with no significant medical history.

### Sample collection

Blood samples were collected immediately prior to starting ETI and then repeated from the first clinical follow-up visit onward, typically occurring at 3 months, but up to one-year post-commencing ETI. This extended window was wide due to the impact of COVID-19 restrictions in our clinic. Some clinical visits were conducted at home and were not suitable for obtaining samples due to potential delays in processing. Samples were collected, processed as per the individual requirement of each analytical technique and stored frozen at −80°C.

### Analytical techniques

A brief overview of the analytical techniques is provided, with more detailed descriptions available in the supplementary methods.

### Peripheral immune cell profiling

Samples were stored with a Cytodelics whole blood cell stabiliser to prevent degradation/cell death during freezing, and subsequently thawed and analysed by flow cytometry.

### Plasma proteomics

Samples were depleted of the top 14 most abundant proteins. Remaining proteins were digested using beads and pepsin, and then analysed by tandem mass spectrometry.

### Cytokine profiling

Plasma samples were analysed using the MSD U-PLEX Biomarker Group 1 (human) 71-Plex kit (Meso Scale Discovery, catalogue # K15081K-1) as per the manufacturer’s instructions.

### Clinical data

All routine clinical data was collected, including spirometry which was performed either in clinic or at home using the Bluetooth® Air Next Spirometer device (Nuvoair, Stockholm, Sweden) (15).

### Statistical and bioinformatic analysis

All data was analysed using R (version 4.1.1). Differences between groups for flow cytometry and immunoassay datasets were tested using t-test or Mann-Whitney U test, depending on the data distribution. Benjamini-Hochberg was applied to the multiplex immunoassay to correct for false discovery. Statistical significance was determined by p-value or adjusted p-value, with threshold 0.05. Proteomics data analyses are detailed within the supplementary methods. In brief, data was processed including steps for normalisation via variance stabilisation normalisation (VSN) (16), batch correction via Combat (17) and removal of proteins with more than 20% of values missing using package missForest (18). Differential expression (DE) analysis was performed using the limma package. Differentially expressed proteins were identified using a Benjamini-Hochberg false discovery rate (FDR) threshold of 0.05. Protein Set Enrichment Analyses (PSEA) were undertaken to evaluate pathway enrichment of expressed proteins between groups using the Fast Gene Set Enrichment Analysis package (19) on a variety of databases via clusterProfiler (20). All code used for proteomics analyses, including packages, functions and concrete parameters used is available in public GitHub repository https://github.com/CBFLivUni/preprocessing_plasma_SPRINT.

## RESULTS

### Clinical and demographic characteristics

We obtained samples pre- and post-commencing ETI in 57 CF subjects (Supplementary Figure E1). Subjects receiving either dual modulators at baseline or who were treatment naïve were combined into one cohort, as there were few differences observed in our datasets when stratified by baseline modulator status (Supplementary Figure E2-E5). Table 1 presents clinical and demographic characteristics for the CF subjects and the 29 healthy controls. The two cohorts were well balanced for age and gender. Individual characteristics are shown within the supplementary material (Supplementary Table E2). The mean time interval between baseline and for repeat sampling on ETI was 133 days (SD = 67). The mean (SD) FEV_1_% increased between starting ETI and the time of repeat sampling (66.0(18.2)% vs 77.9(21.7)%, p=0.002).

**Table 1.**
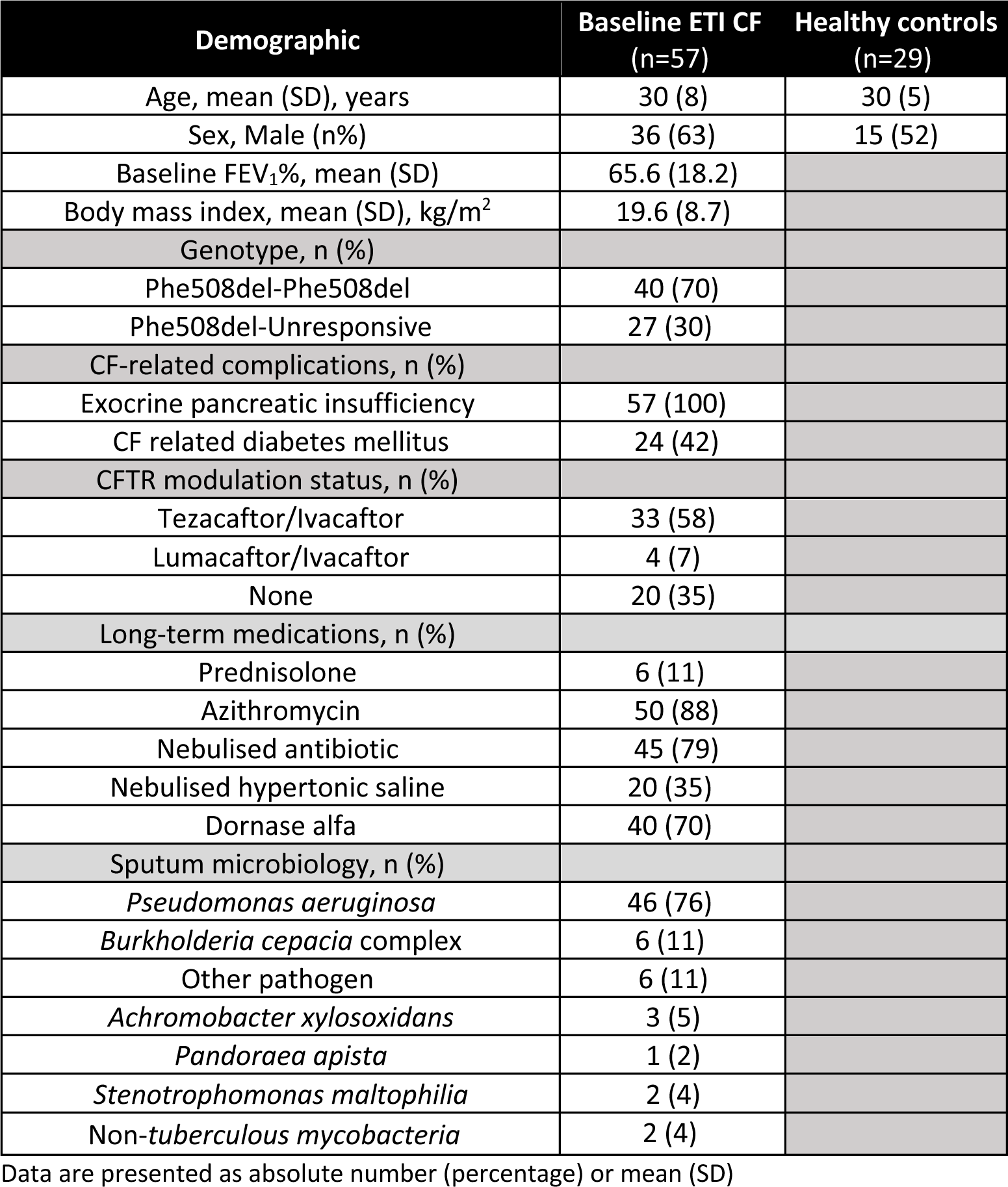
Subject demographic and clinical characteristics.

### The effect of ETI on immune cell populations

To determine immune cell profiles in healthy versus CF subjects, and the effect of ETI therapy on immune cells, we used high-dimensional flow cytometry to identify different innate and adaptive immune populations. Higher total numbers of immune cells (CD45+) were observed in CF patients pre-ETI compared to healthy controls, primarily due to increased levels of circulating neutrophils and monocytes (Figure 1A-B). Further analyses indicated these monocyte differences were due to increased numbers of the classical (CD14+) and intermediate (CD14+ CD16+) subsets (Figure 1C). There were also higher numbers of eosinophils and basophils, but not in the adaptive immune cell populations (Supplementary Figure E6).

**Figure 1.**
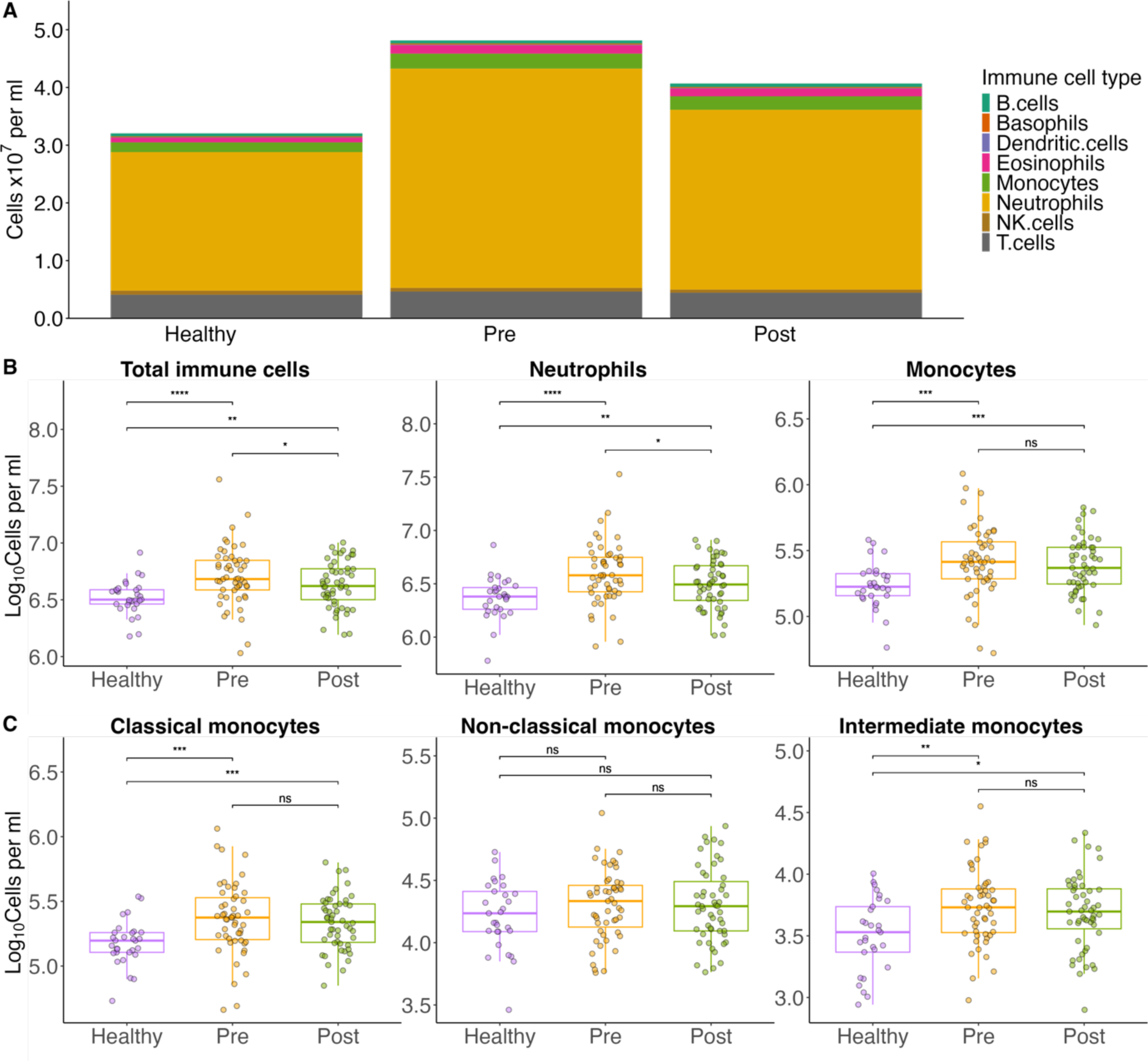
Quantitative profiling of circulating immune cells pre- and post-ETI. Absolute cell counts were measured by high-dimensional flow cytometry. (A) Stacked bar chart displaying median values of selected circulating immune cells. (B) Box plots of the selected immune cells frequently implicated in CF lung disease pathology, with CF subjects stratified according to ETI status. (C) Box plots of the monocyte subsets. For each boxplot, individual cell count, together with median and interquartile ranges are plotted. Difference between cohorts are calculated by t-tests: **** = p < 0.0001, *** = p < 0.001, ** = p < 0.01, * = p < 0.05

With ETI therapy, neutrophil levels reduced, leading to an overall reduction in the number of immune cells (Figure 1A-B). However, ETI therapy did not lead to changes in other immune cell types (Figure 1A-C, Supplementary Figure E6). The post-ETI immune profile remained distinct from healthy controls, with persistently higher levels of neutrophils and monocytes (Figure 1B). Together, these results indicate that ETI does not normalise the systemic immune cell profile in CF patients.

### The effect of ETI therapy on neutrophil and monocyte phenotypes

Although levels of neutrophil and monocytes remained elevated post-ETI therapy, treatment could potentially affect the functionality of these immune cells (Figure 2A-D). We observed that prior to ETI therapy, neutrophil populations in comparison to healthy controls showed reduced expression of CD15, a cell marker that reflects neutrophil maturation within the bone marrow (Figure 2A) (21). With ETI therapy, there was a shift toward a more mature phenotype with increased CD15 expression, as well as other markers related to neutrophil maturation, such as CD16 and CD101 (22). Post-ETI CD15 expression was no longer significantly different to heathy controls. There was a significant increase with ETI in CD206 expression on neutrophils (Figure2B) and monocyte subsets (Supplementary Figures E7-9), surpassing levels observed in healthy controls. Notably, pre-ETI CD206 expression in neutrophils and monocytes was comparable to healthy controls. CD206 expression was previously described on macrophages, and more latterly on neutrophils and monocytes, and is associated with immune resolution and wound repair, consistent with a more anti-inflammatory phenotype (23–26).

**Figure 2.**
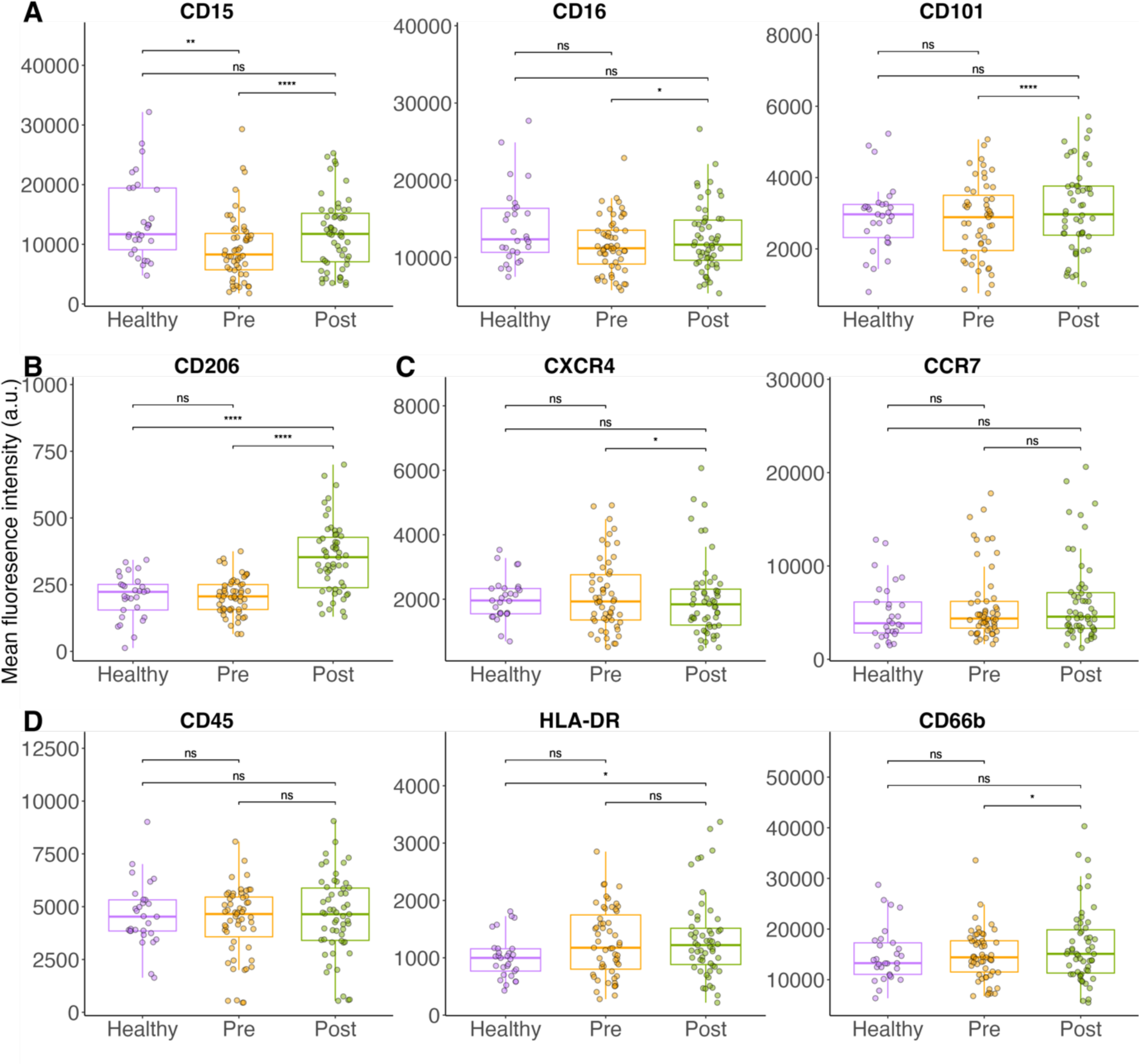
Immunophenotyping of neutrophils pre- and post-ETI. Median fluorescence intensity (MFI) was measured by high-dimensional flow cytometry. MFI values are plotted, with CF subjects stratified according to ETI status. For each group, individual median cell marker MFI, together with median and interquartile ranges are plotted. Difference between cohorts are calculated by Mann Whitney: **** = p < 0.0001, *** = p < 0.001, ** = p < 0.01, * = p < 0.05.

The effect of ETI was also explored on monocyte phenotypes (supplementary figure E7-E8). Specifically, we focussed on classical and intermediate monocytes subsets, as these were elevated pre-ETI therapy compared to healthy controls. There were consistent changes in both classical and intermediate subsets with increased CD101 and HLA-DR expression. Monocytes expressing CD101 have also been associated with beneficial immunomodulatory effects consistent with the immunophenotype shift relating to CD206 expression (27). In contrast, reduced expression of HLA-DR on monocytes is associated with detrimental immunosuppression in conditions such as sepsis (28). As such, the increased levels of HLA-DR suggest a positive effect on immune function.

### The effect of ETI therapy on soluble immune mediator profiles

Cytokines and chemokines, collectively referred to as soluble immune mediators, influence the levels and functionality of circulating immune cells. We explored the effect of ETI therapy on the levels of 61 soluble mediators (Supplementary Table E3) using multi-plex immunoassays. There were 16 that differed between CF subjects and healthy controls (adjusted p-value < 0.5), with 15 showing increased levels (Figure 3A, Supplementary Figure E9). Some of these soluble mediators, such as interleukin 8 (IL8), IL6 and granulocyte-colony stimulating factor (GCSF), have well-established effects on the levels and immunophenotype of circulating neutrophils (29–31), while others, such as macrophage colony-stimulating factor (M-CSF) and C-C motif chemokine ligand 2 (CCL2), primarily influence monocyte biology (32). After ETI treatment, levels of six soluble mediators - IL6, IL8, G-CSF, IL3, C-X-C motif chemokine ligands 1 and 5 (CXCL1 and CXCL5) - significantly reduced, all of which are known to influence levels of circulating neutrophils by a range of mechanisms (Figure 3B-C)(33, 34). There were no differences before commencing ETI in the levels of IL3, CXCL1 or CXCL5 in pwCF compared to healthy controls. Notably, while IL-6 levels reduced with ETI, they remained significantly elevated compared to healthy controls. Most other soluble mediators that were elevated in CF showed minimal response to ETI therapy with 9 remaining significantly different from healthy controls, including M-CSF and CCL2 (Supplementary Figure E10). These changes were not visible within principal component analysis (Supplementary Figure E11)

**Figure 3.**
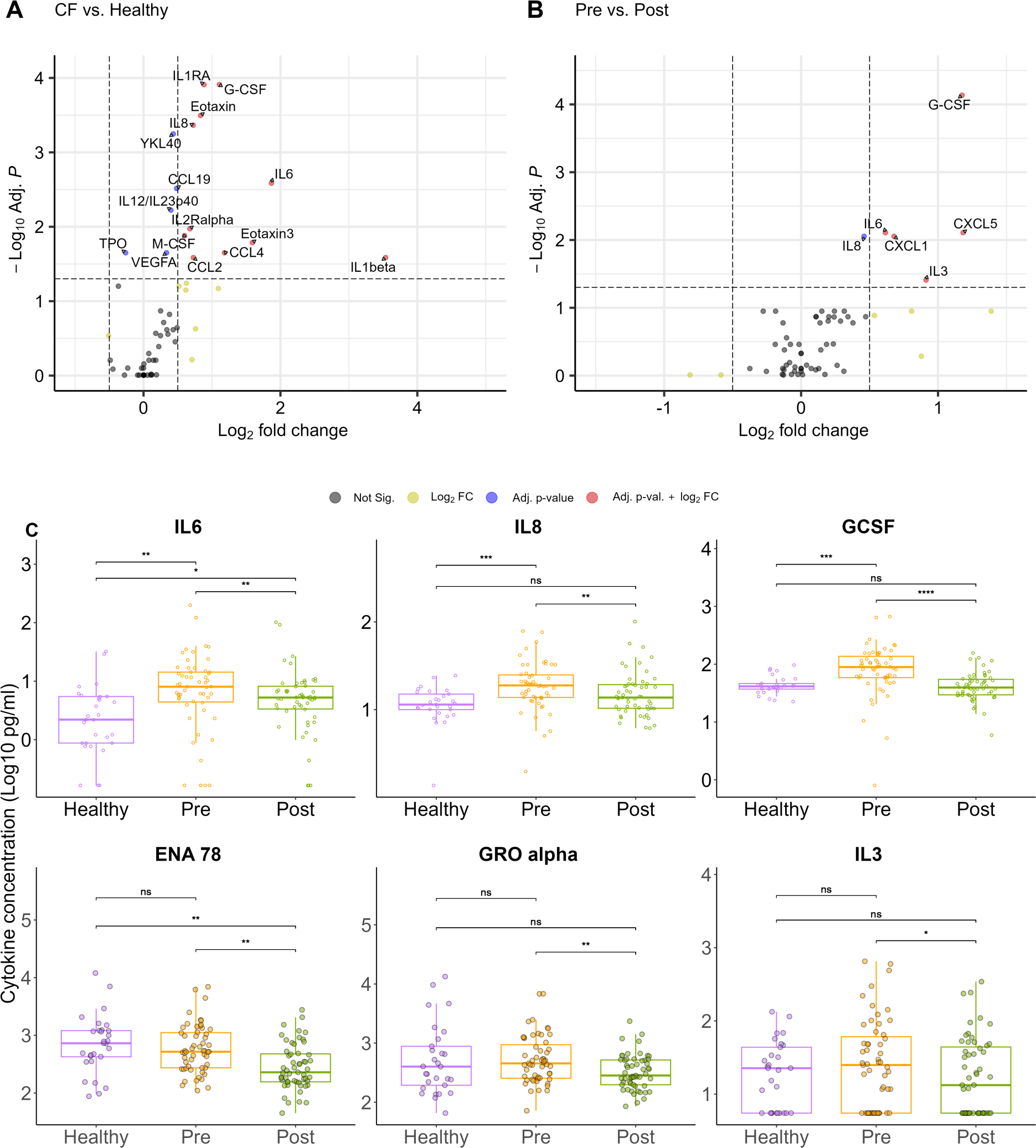
Quantitative profiling of systemic soluble immune mediators pre- and post-ETI. (A) Volcano plot displaying pre-ETI CF versus healthy controls with log2median fold change in soluble mediator abundance against adjusted p-value calculated by Mann-Whitney U tests. Dots are coloured by thresholds based on log-fold change and adjusted p value. Horizontal dashed line indicates cut off for adjusted p value < 0.05. Vertical dashed lined indicates positive and negative cut offs for absolute log2 fold change of 1. (B) Volcano plot showing pre- versus post-ETI. (C) Box plots of key altered soluble mediators. For those that were different at baseline between healthy and pre-ETI and then changed with therapy (adjusted p-value <0.05) the absolute concentrations are shown, with CF subjects stratified by ETI state. For each group, individual median cell marker MFI, together with median and interquartile ranges are plotted. Difference between cohorts are calculated by Mann Whitney with adjustment for false discovery rate: **** = p < 0.0001, *** = p < 0.001, ** = p < 0.01, * = p < 0.05.

The effect of genotype and duration of therapy on the ETI response was explored across all datasets for those variables that changed with ETI therapy. This included variables from the immune cell counts, immune cell markers and soluble mediators. We did not identify any significant influence of genotype (Supplementary Table E4) or duration of ETI therapy on the changes observed in these variables (Supplementary Table E5).

### The effect of ETI on the plasma proteome

Next, we investigated differences in the plasma proteome. This was complementary to soluble mediator profiling, as many cytokines or chemokines are too low abundance to be detected by plasma proteomics. A total of 426 proteins were identified, meeting the criteria of at least two unique peptides and a false discovery rate (FDR) of less than 1%. There were only four differentially expressed proteins between CF and healthy controls, these were Inter-alpha-trypsin inhibitor heavy chain H2 (ITIH2), Leucine rich alpha-2-glycoprotein 1 (LRG1), Peroxisomal membrane protein11B (PEX11B) and Secreted phosphoprotein 2 (SPP2) (Figure 4A and 4C). There was no differential protein expression with ETI therapy (Figure 4B). To gain further function insights we undertook PSEA. This is a ranked overrepresentation method that tries to find significant pathways/functions that differ between two conditions in this case the fold change between CF subjects and healthy controls, and then for the effect of ETI therapy (pre vs post). Importantly, in contrast to other enrichment approaches PSEA utilises the entire list of proteins rather than only those that have been identified *a priori* based upon a statistical threshold. Results showed a significant overrepresentation of innate immune proteins, including those related to neutrophil degranulation in CF patients pre-ETI compared to healthy controls (Figure 4D). The individual proteins responsible for up-regulating these enrichment signals were identified (Supplementary Table E5). These included several proteins with well-described roles in CF inflammation, such as calprotectin (S100A8/S100A9) and C-reactive protein (CRP) (35). With ETI there was decreased expression of innate immune proteins, particularly within the complement pathway (Figure 4E). Those proteins responsible for down-regulating these enrichment signals were identified (Supplementary Table E6) and included CRP and calprotectin. These changes were not visible within principal component analysis (Supplementary Figure E12).

**Figure 4.**
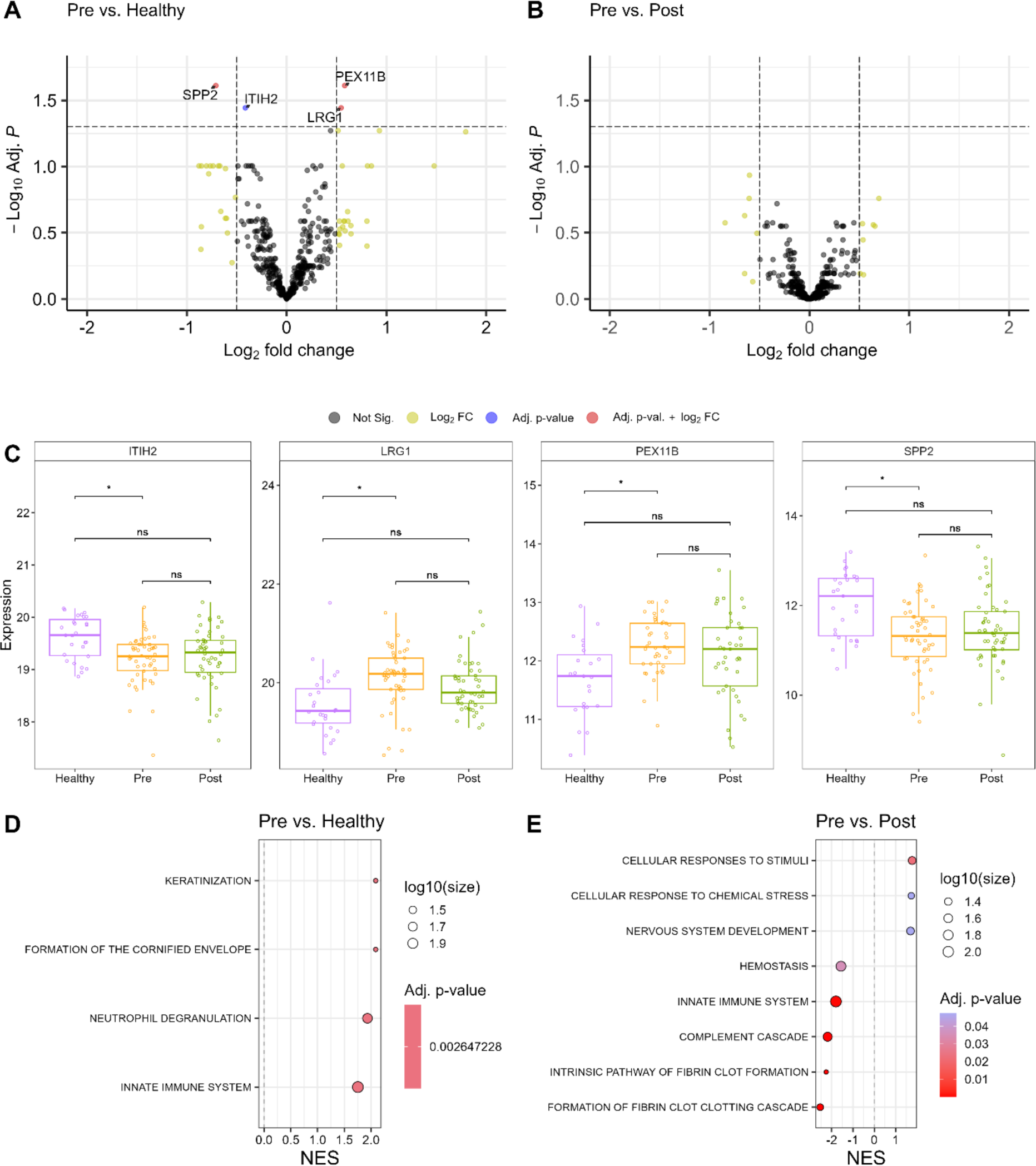
Quantitative profiling of cystic fibrosis plasma proteomes pre- and post ETI. (A) Volcano plot for pre-ETI CF versus healthy displaying fold change as calculated by limma against adjusted p-values. Dots are coloured by thresholds based on log-fold change and adjusted p value. Horizontal dashed line indicates cut off for adjusted p value < 0.05. Vertical dashed lined indicates positive and negative cut offs for absolute log2 fold change of 1. (B) Volcano plot of pre- versus post-ETI CF. (C) Box plots of differentially expressed proteins. For those that were different at baseline between healthy and pre-ETI (adjusted p-value <0.05) the absolute concentrations are shown, with CF subjects stratified by ETI state. For each group, individual median cell marker MFI, together with median and interquartile ranges are plotted. Difference between cohorts are calculated by limma with adjustment for false discovery rate: * = p < 0.05. (D) The protein set enrichment analysis for CF vs healthy. X-axis indicates the Net Enrichment Scores (NES) of a pathway, with a negative score indicating that the pathway was downregulated in the CF group compared to the healthy group and a positive score indicating that the pathway was upregulated. The colour of the dot reflects p adjusted by Benjamini-Hochberg correction. The size of the dot represents the number of proteins present in the pathway that were also present in the data. (E) Protein set enrichment analysis of pre- versus post-ETI therapy. Definitions of abbreviations: ITIH2 = Inter-alpha-trysin inhibitor heavy chain H2, LRG1 = Leucine rich alpha-2-glycoprotein 1, PEX11B = peroxisomal membrane protein11B, SPP2 = Secreted phosphoprotein 2.

### The relationship between ETI-related changes in systemic inflammation and lung function

Finally, we investigated the relationship between changes with ETI therapy observed for systemic inflammation with changes in lung function. Given the neutrophilic nature of the effects observed with ETI, we focussed on this signature in the cell counts, cell markers and soluble mediators and then only on those variables that showed a significant change with therapy (Supplementary Table E7). There were only inverse correlations found between changes in two of the neutrophil cell markers – CXCR4 and CD16 - and lung function. As would be expected if these changes were influencing lung function, CXCR4 expression significantly decreased with ETI therapy, whereas in contrast CD16 expression increased with ETI. Furthermore, there were no differences between pre-ETI and healthy for either neutrophil cell marker.

## DISCUSSION

In this study, we determined the effects of ETI on systemic inflammation in PwCF with at least one Phe508del allele. We observed prior to commencing ETI therapy that the immune profile was characterised predominantly by excessive neutrophilic inflammation. This excessive neutrophilic inflammation whilst reducing with ETI did not fully resolve. There were lower levels of circulating neutrophils, but these did not normalise with therapy. In contrast monocytes appeared unaffected and thus also remained elevated. Despite levels of immune cells remaining elevated in post-ETI CF subjects there was a shift toward a less inflammatory immunophenotype for both neutrophils and monocytes. Changes were also observed in the soluble immune mediator profile and plasma proteome, with levels of six inflammatory immune mediators reduced, and down-regulation of the innate immune and the complement protein sets. Finally, although we identified weak associations between the changes in the expression of two neutrophil cell markers and lung function, the pre-therapy expression of these markers relative to healthy controls, and the changes associated with ETI therapy did not suggest a plausible direct influence on CF lung disease.

It is well-established that PwCF prior to commencing ETI therapy have excessive systemic neutrophilic inflammation (10, 11). Accordingly, we observed immune cell profiles that in CF subjects in comparison to healthy controls had elevated levels of circulating neutrophils and expression of cell markers consistent with a less mature, more inflammatory phenotype. Consistent with previous work the levels of circulating neutrophil levels fell with ETI therapy (10, 36). However, in contrast to these studies, we demonstrate that circulating neutrophil levels do not normalise. This likely reflects the strength of our comparisons between suitably large cohorts of CF subjects commencing ETI and age-matched healthy controls. Although Dhote and colleagues measured immune cell change over a longer period of ETI therapy, we do not believe this solely accounts for the normalisation of neutrophil counts they describe, as we observed no significant relationship between time on therapy and neutrophil counts. Instead, this likely reflects our study making comparisons with prospective age-matched healthy controls rather than against clinical reference ranges.

The neutrophil immunophenotypes revealed several novel findings about the influence of ETI therapy on systemic inflammation. We observed a shift toward a more mature neutrophil phenotype with therapy, characterised by increased expression of CD15, CD16 and CD101. Additionally, CD206 expression increased to levels significantly higher than those observed in healthy controls, consistent with a transition to an immune-resolving phenotype (25, 26). Notably, although monocyte levels did not change with ETI, there were important changes in their phenotype. There was a similar shift in CD206 expression in monocytes as that observed on neutrophils, that may also reflect an immune resolving phenotype. This increased expression has only previously been described in CF for cultured and stimulated monocytes (37).

Persistent neutrophilic inflammation has the potential to adversely influence long-term outcomes for PwCF. Neutrophils are highly toxic cells and damage tissues by multiple mechanisms, including by releasing proteases and neutrophil extracellular traps (38). Although, damage might be confined to one organ, such as the lung where inflammation has been shown to persist after ETI, it is also possible that there could be more systemic consequences. Chronic systemic inflammation has been associated with an increased risk of developing a range of other conditions including cardiovascular disease, malignancy and neuro-degenerative conditions (9). However, the changes we observed in immunophenotype with ETI might mitigate some of the potential neutrophil-related toxicity. CF neutrophils are a heterogeneous population of immune cells, with different subsets exhibiting distinct phenotypic and functional characteristics (39). The shift observed toward a more mature phenotype would result in less active neutrophils that have a shorter lifespan, whereas the immune-resolving phenotypes of neutrophils and monocytes could also further reduce neutrophil activity. The increased CD206 expression seen with ETI on both neutrophils and monocytes is particularly intriguing. CD206 is a mannose receptor, typically associated with alternatively activated macrophages of M2-like phenotype (23, 24). However, there is increasing evidence that CD206 is also an important marker of neutrophil and monocyte function, suggesting that an immune resolving state may still be ongoing when sampling was repeated on therapy (25, 26). Further study is required to better understand the role of these immune cell phenotypes in both local and systemic inflammation in PwCF, as well as how they might impact long-term clinical outcome.

The multi-modal and multi-dimensional assessment of systemic inflammation undertaken here has allowed the soluble immune mediator profile and plasma proteome to also be characterised, and then further interrogated for the effects of ETI therapy. We observed increased levels of soluble mediators that have been previously implicated in CF lung disease pathogenesis, including IL6, IL8, G-GSF and IL1-beta (40). However, there were also differences seen in other lesser-studied blood soluble mediators, such as those relating to Th2-mediated inflammation (Eotaxin and Eotaxin-2) and regulating circulating monocyte levels (M-CSF and CCL2). There were six soluble mediators that reduced with ETI, all known to influence circulating neutrophil levels. Of these changes in IL3 and G-CSF have not been described previously. Notably, amongst the eight soluble mediators that remained persistently elevated after ETI therapy were M-CSF and CCL2, which is also a novel finding.

We revealed by PSEA that innate immune proteins were overexpressed in PwCF, which includes within this protein set many that are categorised as related to neutrophil degranulation. Elevated levels of some innate immune proteins, such as calprotectin and C reactive protein (CRP), have been well-described in CF populations. With ETI therapy innate immune protein expression reduced, including within the complement system protein set. Although complement activity has been implicated in CF lung disease pathology, it’s response to ETI therapy has not been previously described.

Our soluble immune mediator profiling suggests that ETI does not uniformly influence all aspects of systemic inflammation. The soluble mediators that influence circulating neutrophil levels reduce with ETI therapy, whereas those influencing monocyte levels appear unchanged. The importance of these elevated monocytes and their regulatory mediators requires further investigation, including how they might affect patients established on ETI therapy. This untreated aspect of systemic inflammation may represent a potential therapeutic target. Similarly, the role of complement in CF requires additional exploration as therapies reducing its activity may also have utility.

Finally, we failed to identify any systemic immune signatures that correlated with the improvements in lung function observed with ETI therapy. This may reflect that it is airway inflammation, rather than systemic inflammation, directly influences lung function. Therefore, the next crucial step is to gain a more comprehensive understanding of the relationship between systemic inflammation, airway inflammation and CF lung disease. This is an important goal, as identifying specific systemic immune mechanisms related to airway inflammation may produce novel therapeutic targets or potential surrogate measures of airway inflammation.

This study has encountered several notable limitations. There was unavoidable variation in the time between initiating ETI and repeat sampling, caused by COVID-related restrictions on clinical contact for PwCF, and represents another potential confounder. However, when tested for we found no influence of timing of sampling upon our results. Unfortunately, we were not able to assess airway inflammation as most participants were unable to produce spontaneous sputum once exposed to ETI. In those that did produce sputum many were unpaired with blood sampling. Further studies seeking to validate our finding in patients established on ETI therapy should ideally attempt simultaneous assessment of systemic and airway inflammation, potentially obtaining respiratory samples from sputum induction or bronchoscopy with lavage. Although, plasma proteomics offered valuable insights, including highlighting the potential importance of complement activity, insufficient subject numbers hindered the full utilisation of this approach. Our findings suggest that much of the proteome is not influenced by CF and there is significant overlap between disease and healthy protein levels. As such, future studies might be more successful identifying individual proteins with larger cohort sizes and by taking a targeted approach, e.g. focussing on proteins related to innate immunity.

In conclusion, whilst ETI reduced excessive systemic inflammation in PwCF it did not fully resolve. The most prominent improvements were within the levels and phenotypes of circulating neutrophils, as well as in the soluble immune mediators that regulate their activity. However, there was no clear relationship between ETI-related changes in systemic inflammation and lung function. The long-term consequences of persisting systemic inflammation after ETI therapy needs to be established for PwCF, in order to identify potentially adverse effects on organ systems and direct future therapeutic interventions.

## Data Availability

All data produced will be made available once the manuscript is published in a peer-reviewed journal

## ACKNOWLEDGEMENTS

This report is independent research supported by the North West Lung Centre Charity at Manchester University NHS Foundation Trust. The views expressed in this publication are those of the author(s) and not necessarily those of the NHS, the North West Lung Centre Charity or the Department of Health. The authors would like to acknowledge the Manchester Allergy, Respiratory and Thoracic Surgery Biobank, the North West Lung Centre Charity and the National Institute for Health Research (NIHR) Manchester Clinical Research Facility for supporting this project. In addition, we would to thank the study participants for their contribution, as well as the medical and nursing staff especially Jo Hyde, Cassandra McNaughton and Daniel Tewkesbury at the Manchester Adult CF Centre for their invaluable assistance. Professor Smith is funded by the Manchester NIHR BRC and a Wellcome Investigator Award.

