## Supplementary material for "The effect of highly effective modulator therapy on systemic inflammation in cystic fibrosis"

**Financial support:** This research was funded by a project grant from the North West Lung Centre Charity.

### Supplementary methods

#### Flow cytometry

For all donors, 1 ml blood was mixed with 1 ml Cytodelics whole blood cell stabiliser (Cytodelics AB, Sweden). Samples were incubated at room temperature (RT) for 10 minutes and then stored at -80°C. The Cytodelics whole blood stabiliser allows preservation of granulocytes following -80°C storage and is suitable for flow cytometric analysis of cell surface antigens. Thawing, red blood cell lysis and washing steps were performed according to the manufacture's description prior to staining for flow cytometry analysis.

To assure accurate immune cell count quantification, prior to antibody staining 25 µl of known concentration CountBright Absolute Counting Beads (Molecular Probes, USA) were added to each sample. To block unspecific staining, samples were suspended in human TruStain FcX blocking solution (Biolegend, USA) and incubated for 20 minutes at RT. Subsequently, an antibody cocktail for cell surface markers was added (Supplementary Table E1) and incubated in the dark for 30 minutes at RT. After washing with phosphate-buffered saline containing 2% fetal calf serum and 2 mM EDTA, samples were acquired using BD FACSymphony A5 cell analyser with BD FACSDiva software version v9.0 (BD Biosciences, USA). Obtained flow cytometry data were analysed with Flowjo software v.10.10 (BD Biosciences, USA) and statistical analysis performed using Prism 10 software (GraphPad Software Inc, USA).

#### Plasma proteomics

*High Select Top14 protein depletion*

The Thermo Scientific™ High-Select™ Top14 Abundant Protein Depletion Resin (ThermoFisher, A36372) employs antibody-based removal of 14 high-abundant proteins from human plasma: Albumin, IgA, IgD, IgE, IgG, IgG (light chains), IgM, Alpha-1-acid glycoprotein, Alpha-1-antitrypsin, Alpha-2-macroglobulin, Apolipoprotein A1, Fibrinogen, Haptoglobin and Transferrin. An optimised method was employed, combining 5 µL of human plasma with 300 µL of depletion resin into each well of a 96-well lo-bind PCR plate (Eppendorf, 0030129504). The mixture was incubated at RT (21°C) for 15 min with shaking at 1400 rpm before centrifuging through a 96-well filter plate (Agilent, 200957-100) for 2 min at 400 rcf at RT. The flow through was collected in a low-bind PCR plate at a total volume of 150 µL. Finally, SDS

(ThermoFisher, 1281-1680) was added to a final concentration of 1% (v/v) and the plate was heated to 95°C for 10 min without shaking.

*Protein digestion*

From each well, 50µL of depleted plasma was reduced and alkylated by the addition of 10 µl of 60 mM TCEP (ThermoFisher, UI287564) and 240 mM CAA (Sigma-Aldrich, C0267), respectively and heated to 95°C for 5 min. To each well, 10 µL of Sera-Mag SpeedBeads A and B (GE Healthcare, 45152105050250 and 65152105050250), at equal ratio, were added, before the addition of 130 µL of 100% ethanol (v/v) (Honeywell, 34870). After a 10 min incubation at RT, the supernatant was discarded and the beads were washed 6x with 300 µL 80% ethanol (v/v). On-bead digestion was performed by incubating each sample with 300 ng of LysC (info) in 150 µl 100mM ammonium bicarbonate (Sigma-Aldrich, A6141) for 2 hours at 37 °C with shaking at 1400 rpm before the addition of 300 ng of trypsin (Promega, V5280). Samples were incubated overnight (16h) at at 37 °C with shaking at 1400 rpm. Digestion was stopped by the addition of 5 µL of 10% TFA (v/v) (info).

*Proteomic analysis*

A total peptide concentration of 2.5ug per sample was analysed by liquid chromatography mass spectrometry using an Evosep One (Evosep, EV-1000) coupled to an Orbitrap Exploris 480 mass spectrometer (Thermo Fisher Scientific) using the 30 sample per day Evosep liquid chromatography gradient and data independent acquisition (DIA) method consisting of a full scan at FAIMS CV -40, a DIA scan at -40, a full scan at -60, and a DIA scan at -60. Full scans were acquired in the m/z range 345-900 with 120 000 resolution, 300% normalised AGC target, automatically selected max IT, and in profile mode. Data-independent fragments were acquired in 42 fixed-length windows of 13 m/z in the m/z range 350-896 with no overlap, and with window placement optimization. A full scan was acquired after every 14 DIA scans, taking 3 full scans (with each CV) to fragment the ions in all 42 DIA windows. A full duty cycle consists of 6 MS1 scans and 84 MS2 scans. MS2 resolution was set to 15 000, NCE to 32%, AGC target to 1 000%. The m/z range of the fragmented ions and the max IT were both automatic. DIA data was recorded in profile mode.

**Multiplex immunoassays**

Cytokine analysis was performing using the MSD U-PLEX Biomarker Group 1 (human) 71-Plex kit (Meso Scale Discovery, K15081K-1) as per the manufacturer's instructions. Briefly, individual U-PLEX-coupled antibodies were created and used to coat each well of a 96-well plate. Calibrator standards were created and added to assigned wells, remaining wells were filled with 25 µl of plasma. After the addition of a detection antibody solution and read buffer, plates were analysed by using a Methodical Mind software on MESO QuickPlex SQ 120MM (Meso Scale Discovery, AI1AA-0) instrument.

**Statistical methods for plasma proteomics**

*Pre-processing*

Proteins that had more than 20% of values missing were removed from analysis using missForest package (Stekhoven *et al*., 2022). Variance Stabilisation Normalisation (VSN) (version 3.62.0) was performed separately within each plate. VSN requires untransformed data as an input as it includes a process similar to log transformation (Huber *et al.,* 2002). Batch effect was corrected across plates using ComBat from the sva R package (version 3.42.0) (Johnson *et al.,* 2007).

*Statistical Analyses*

All data pre-processing and statistical analyses were completed using R (version 4.1.1). The R package variancePartition (version 1.24.1) was used to establish the covariates that drove differences between pre- and post-ETI, and healthy and pre-ETI protein expression. This involves fitting a linear mixed model to each protein where categorical variables are modelled as random effects and continuous variables are models as fixed effects. Covariates baseline age, baseline FEV1%, baseline weight and time between first and second patient visit showed greater median variance explained across all models fit to each protein than the cohort that samples were part of. These covariates were selected for further analysis.

Differential expression (DE) analysis was performed using limma (version 3.50.3), an approach using linear models and empirical Bayes. Akaike Information Criterion (AIC) was calculated for each model where every combination of covariates being included in the model or not was fit i.e. a model was fit including age, FEV1%, weight and times between visits as covariates, and another model was fit including only age, FEV1%, weight, but not time between visits and so on. The model with the lowest resulting AIC, indicating best model fit, was the model that included no additional covariates. Therefore, no covariates were included for DE analysis. DE analysis was run between pre- vs post- ETI, pre- vs healthy controls, and post- vs healthy controls. DE proteins were identified using a Benjamini-Hochberg false discovery rate (FDR) threshold of 0.05.

*Protein Set Enrichment Analyses*

Proteins were mapped to gene symbols and the Fast Gene Set Enrichment Analysis (fsgea) R package (version 1.20.0) was used to evaluate pathway enrichment of expressed proteins in the pre- vs post- ETI groups using the corresponding gene symbols. Enrichment was calculated for hallmark MSigDB, Wikipathways, Reactome and KEGG gene sets retrieved from MSigDB.

***Table E1.*** **Antibody cocktails for cell surface markers**

| Target | Fluorophore | Clone | Concentration | Supplier | Catalogue No |
|---|---|---|---|---|---|
| CD66b | FITC | G10F5 | 1:100 | Biolegend | 305104 |
| CD11c | PerCP-Cy5.5 | 3.9 | 1:50 | Biolegend | 301624 |
| CD15 | APC | W6D3 | 1:100 | Biolegend | 323008 |
| CD16 | AF700 | 3G8 | 1:100 | Biolegend | 302026 |
| HLA-DR | APC-Cy7 | L243 | 1:50 | Biolegend | 307618 |
| CD206 | BV421 | 15-2 | 1:200 | Biolegend | 321126 |
| CD45 | BV510 | HI30 | 1:50 | Biolegend | 304036 |
| CD193 | BV605 | 5E8 | 1:100 | Biolegend | 310716 |
| CD3 | BV650 | UCHT1 | 1:100 | Biolegend | 300468 |
| CD19 | BV711 | HIB19 | 1:100 | Biolegend | 302246 |
| CD20 | BV711 | 2H7 | 1:100 | Biolegend | 302342 |
| CD123 | BV786 | 6H6 | 1:100 | Biolegend | 306032 |
| CD14 | PE | M5E2 | 1:50 | Biolegend | 301806 |
| CD184 (CXCR4) | PE-CF594 | 12G5 | 1:50 | Biolegend | 306526 |
| CD197 (CCR7) | PE-Cy7 | G043H7 | 1:50 | Biolegend | 353226 |
| CD101 | BUV395 | V7.1 | 1:100 | BD | 747544 |
| CD56 | BUV737 | NCAM16.2 | 1:100 | BD | 612766 |
| CD141 | BUV737 | 1A4 | 1:100 | BD | 741867 |

**Table E2. Individual clinical and demographic characteristics of CF participants**

| ID | Time on therapy | Baseline modulator | Gene1 | Gene2 | Pancreatic status | Ps | BCC | Baseline FEV1 | Baseline FEV1% | V2 FEV1 | V2 FEV1% |
|---|---|---|---|---|---|---|---|---|---|---|---|
| 2227 | 77 | TEZ/IVA | F508del | F508del | Insufficient | Yes | Yes | 2.34 | 64 | 2.74 | 75 |
| 2271 | 238 | LUM/IVA | F508del | F508del | Insufficient | Yes | No | 1.18 | 36 | 1.34 | 41 |
| 2324 | 161 | None | F508del | F508del | Insufficient | Yes | No | 1.64 | 57 | 1.94 | 67 |
| 2340 | 84 | TEZ/IVA | F508del | F508del | Insufficient | Yes | No | 2.51 | 58 | 3.13 | 72 |
| 2420 | 152 | None | F508del | R792X | Insufficient | Yes | No | 1.7 | 61 | 2.05 | 74 |
| 2511 | 61 | LUM/IVA | F508del | F508del | Insufficient | Yes | No | 2.01 | 52 | 2.5 | 64 |
| 2652 | 203 | TEZ/IVA | F508del | F508del | Insufficient | Yes | No | 1.8 | 58 | 1.8 | 58 |
| 2830 | 182 | TEZ/IVA | F508del | F508del | Insufficient | Yes | No | 3.64 | 86 | 3.72 | 88 |
| 2833 | 56 | TEZ/IVA | F508del | F508del | Insufficient | Yes | Yes | 1.26 | 46 | 1.34 | 49 |
| 2888 | 84 | TEZ/IVA | F508del | F508del | Insufficient | No | Yes | 2.48 | 89 | 2.61 | 94 |
| 2899 | 203 | TEZ/IVA | F508del | F508del | Insufficient | Yes | No | 2.86 | 96 | 3.46 | 117 |
| 3011 | 189 | TEZ/IVA | F508del | F508del | Insufficient | Yes | No | 3.65 | 83 | 4.11 | 93 |
| 3033 | 189 | TEZ/IVA | F508del | F508del | Insufficient | Yes | No | 1.56 | 39 | NA | NA |
| 3067 | 112 | None | F508del | 1517T | Insufficient | Yes | No | 3.08 | 86 | 3.66 | 102 |
| 3118 | 229 | TEZ/IVA | F508del | F508del | Insufficient | No | No | 2.79 | 75 | 3.19 | 86 |
| 3125 | 70 | TEZ/IVA | F508del | F508del | Insufficient | Yes | No | 2.77 | 67 | 2.81 | 67 |
| 3170 | 154 | TEZ/IVA | F508del | F508del | Insufficient | Yes | No | 1.29 | 39 | 1.83 | 56 |
| 3280 | 126 | TEZ/IVA | F508del | F508del | Insufficient | Yes | No | 1.89 | 61 | 2.75 | 88 |

| ID | Time on therapy | Baseline modulator | Gene1 | Gene2 | Pancreatic status | Ps | BCC | Baseline FEV1 | Baseline FEV1% | V2 FEV1 | V2 FEV1% |
|---|---|---|---|---|---|---|---|---|---|---|---|
| 3405 | 96 | TEZ/IVA | F508del | F508del | Insufficient | Yes | No | 1.92 | 50 | 2.54 | 66 |
| 3452 | 259 | None | F508del | 1154insTC | Insufficient | Yes | No | 1.9 | 44 | 2.51 | 59 |
| 3599 | 112 | TEZ/IVA | F508del | F508del | Insufficient | Yes | No | 2.47 | 71 | 2.84 | 82 |
| 4642 | 63 | TEZ/IVA | F508del | F508del | Insufficient | Yes | No | 2.75 | 68 | 3.49 | 86 |
| 4763 | 105 | None | F508del | 2711delT | Insufficient | No | Yes | 2.2 | 54 | 3.08 | 76 |
| 5076 | 196 | None | F508del | 117-1G-A | Insufficient | Yes | No | 3.09 | 89 | 3.68 | 106 |
| 5116 | 112 | TEZ/IVA | F508del | F508del | Insufficient | No | No | 2.87 | 64 | 3.41 | 76 |
| 5117 | 63 | TEZ/IVA | F508del | F508del | Insufficient | Yes | Yes | 2.77 | 68 | 3.42 | 85 |
| 5181 | 63 | None | F508del | R560T | Insufficient | No | No | 2.5 | 65 | NA | NA |
| 5241 | 216 | TEZ/IVA | F508del | F508del | Insufficient | No | No | 2.29 | 55 | 3.74 | 91 |
| 5250 | 63 | None | F508del | /621+1 | Insufficient | Yes | No | 3.2 | 64 | 3.64 | 73 |
| 5268 | 147 | TEZ/IVA | F508del | F508del | Insufficient | Yes | No | 2.19 | 58 | 2.7 | 72 |
| 5270 | 70 | TEZ/IVA | F508del | F508del | Insufficient | Yes | No | 1.79 | 59 | 1.52 | 50 |
| 5369 | 112 | TEZ/IVA | F508del | F508del | Insufficient | Yes | No | 3.59 | 102 | 4.04 | 115 |
| 6013 | 201 | LUM/IVA | F508del | F508del | Insufficient | Yes | No | 2.62 | 77 | 3.06 | 91 |
| 6014 | 70 | TEZ/IVA | F508del | F508del | Insufficient | Yes | No | 4.03 | 92 | 4.5 | 103 |
| 6015 | 87 | TEZ/IVA | F508del | F508del | Insufficient | Yes | No | 1.59 | 35 | 2.4 | 53 |
| 6017 | 126 | None | F508del | N1303K | Insufficient | No | No | 2 | 47 | 2.33 | 55 |
| 6018 | 135 | None | F508del | R553X | Insufficient | Yes | No | 1.84 | 65 | NA | NA |

| ID | Time on therapy | Baseline modulator | Gene1 | Gene2 | Pancreatic status | Ps | BCC | Baseline FEV1 | Baseline FEV1% | V2 FEV1 | V2 FEV1% |
|---|---|---|---|---|---|---|---|---|---|---|---|
| 6020 | 189 | TEZ/IVA | F508del | F508del | Insufficient | No | Yes | 2.88 | 90 | 3.94 | 124 |
| 6021 | 205 | None | F508del | 3659DelC | Insufficient | Yes | No | 1.6 | 54 | 1.49 | 50 |
| 6052 | 76 | TEZ/IVA | F508del | F508del | Insufficient | No | No | 2.6 | 74 | 2.7 | 76 |
| 6053 | 348 | TEZ/IVA | F508del | F508del | Insufficient | Yes | No | 2.18 | 53 | 2.28 | 55 |
| 6054 | 84 | None | F508del | 2184A | Insufficient | Yes | No | 1.35 | 33 | 1.72 | 42 |
| 6057 | 156 | None | F508del | 3007ΔG | Insufficient | Yes | No | 2.43 | 53 | NA | NA |
| 6058 | 155 | None | F508del | R560T | Insufficient | No | No | 3.96 | 85 | 4.54 | 97 |
| 6059 | 126 | TEZ/IVA | F508del | F508del | Insufficient | Yes | No | NA | NA | NA | NA |
| 6060 | 56 | None | F508del | G542X | Insufficient | Yes | No | 3.55 | 69 | 4.07 | 79 |
| 6063 | 58 | TEZ/IVA | F508del | F508del | Insufficient | Yes | No | 1.88 | 48 | 3.21 | 83 |
| 6064 | 91 | None | F508del | c1766+1G>A | Insufficient | Yes | No | 2.86 | 53 | 3.79 | 71 |
| 6066 | 91 | LUM/IVA | F508del | F508del | Insufficient | Yes | No | 0.87 | 32 | 1.1 | 40 |
| 6067 | 322 | TEZ/IVA | F508del | F508del | Insufficient | Yes | No | 2.52 | 54 | 2.94 | 63 |
| 6091 | 63 | None | F508del | R553X | Insufficient | Yes | No | 2.98 | 79 | 4.21 | 112 |
| 6092 | 168 | TEZ/IVA | F508del | F508del | Insufficient | Yes | No | 2.64 | 77 | NA | NA |
| 6127 | 134 | None | F508del | W1098R | Insufficient | Yes | No | 3.25 | 74 | 4.39 | 100 |
| 6128 | 105 | TEZ/IVA | F508del | F508del | Insufficient | Yes | No | 2.51 | 55 | 3.11 | 68 |
| 6130 | 91 | None | F508del | F508del | Insufficient | Yes | No | 2.78 | 72 | NA | NA |
| 6131 | 91 | None | F508del | F508del | Insufficient | Yes | No | 3.19 | 76 | 3.72 | 90 |

| ID | Time on therapy | Baseline modulator | Gene1 | Gene2 | Pancreatic status | Ps | BCC | Baseline FEV1 | Baseline FEV1% | V2 FEV1 | V2 FEV1% |
|---|---|---|---|---|---|---|---|---|---|---|---|
| 6139 | 84 | TEZ/IVA | F508del | F508del | Insufficient | No | No | 3.68 | 82 | 4.13 | 93 |

Abbreviations: Pseudomonas aeruginosa (Ps), Burkholderia cepacia complex (Bcc), Percent predicted FEV1 (FEV1%), M (male), F(female), ivacaftor (IVA), lumacaftor (LUM), tezacaftor (TEZ), not available data (NA)

**Table E3. U-PLEX Biomarker Group 1 (human) 71-Plex**

| | | | | | | |
|---|---|---|---|---|---|---|
| CTACK | I-309 | IL-5 | IL-17A | IL-27* | MIF | TRAIL |
| ENA-78 | IFN-α2a | IL-6 | IL-17A/F | IL-29/IFN-λ1* | MIP-1α | TSLP* |
| Eotaxin | IFN-β | IL-7 | IL-17B | IL-31* | MIP-1β | VEGF-A |
| Eotaxin-2 | IFN-γ | IL-8 | IL-17C | IL-33* | MIP-3α | YKL-40 |
| Eotaxin-3 | IL-1α | IL-9 | IL-17D | IP-10 | MIP-3β | M-CSF |
| EPO | IL-1β | IL-10 | IL-17E/IL-25* | I-TAC | MIP-5 | |
| FLT3L | IL-1RA | IL-12/IL-23p40 | IL-17F* | MCP-1 | SDF-1α | |
| Fractalkine | IL-2 | IL-12p70 | IL-18 | MCP-2 | TARC | |
| G-CSF | IL-2Rα | IL-13 | IL-21* | MCP-3 | TNF-α | |
| GM-CSF | IL-3 | IL-15 | IL-22* | MCP-4 | TNF-β | |
| GRO-α | IL-4 | IL-16 | IL-23* | MDC | TPO | |

***Plate failed**

### Table E4. Change in selected variables with ETI therapy stratified by genotype

| Dataset | Variable | Change for F508del homozygote (median, IQR) | Change for F508del / minimal function | Fold difference | P value |
|---|---|---|---|---|---|
| Immune cells (cells/mm3) | Neutrophils | -761,166 (-2,182,950 to 898,112) | -1,641,397 (-3,170,437 to -427,365) | 0.46 | 0.09 |
| Neutrophil cell markers (MFI) | CD206 | 127 (76.00 to 214.75) | 101 (62.00 to 156.00) | 0.81 | 0.21 |
| Neutrophil cell markers (MFI) | CD15 | 1750 (-702.25 to 4484.50) | 1524 1011.00 to 3760.00) | 0.87 | 0.40 |
| Neutrophil cell markers (MFI) | CD16 | 661 (-555.00 to 1754.00) | 1105 (-257.00 to 1669.00) | 1.70 | 0.59 |
| Neutrophil cell markers (MFI) | CD101 | 246 (15.50 to 557.25) | 327 (21.00 to 581.00) | 1.33 | 0.36 |
| Neutrophil cell markers (MFI) | CD66b | 1266 (-1866.75 to 3756.25) | 0 (-781.00 1068.00) | NA | 0.87 |
| Neutrophil cell markers (MFI) | CXCR4 | -108 (-321.25 to 102.50) | 175 (-274.00 to 161.00) | 0.67 | 0.80 |
| Cytokines (pg/ml) | IL6 | -3.12 (-8.47 to 1.39) | -5.28 ( -14.21 to -0.50) | 1.70 | 0.19 |
| Cytokines (pg/ml) | IL8 | -4.76 ( -9.48 to 1.29) | -7.03 ( -18.01 to 3.26) | 1.50 | 0.12 |
| Cytokines (pg/ml) | CXCL1 | -81.65 (-350.92 to 69.60) | -118.11 (-241.29 to -48.52) | 1.45 | 0.24 |
| Cytokines (pg/ml) | CXCL5 | -240.69 (-654.29 to 103.52) | -455.45 (-760.73 to -137.37) | 1.89 | 0.30 |
| Cytokines (pg/ml) | G-CSF | -35.37 (-86.37 to -12.91) | -45.56 (-87.68 to -23.50) | 1.3 | 0.40 |

### Table E5. Correlation between change with ETI therapy and duration on therapy

| Dataset | Variable | R correction | P value |
|---|---|---|---|
| Immune cells (cells/mm3) | Neutrophils | -0.236 | 0.096 |
| Neutrophil cell markers (MFI) | CD206 | 0.024 | 0.860 |
| Neutrophil cell markers (MFI) | CD15 | 0.188 | 0.169 |
| Neutrophil cell markers (MFI) | CD16 | 0.156 | 0.255 |
| Neutrophil cell markers (MFI) | CD101 | 0.086 | 0.547 |
| Neutrophil cell markers (MFI) | CD66b | 0.251 | 0.064 |
| Neutrophil cell markers (MFI) | CXCR4 | -0.199 | 0.146 |
| Cytokines (pg/ml) | IL6 | -0.194 | 0.160 |
| Cytokines (pg/ml) | IL8 | -0.122 | 0.380 |
| Cytokines (pg/ml) | CXCL1 | -0.058 | 0.675 |
| Cytokines (pg/ml) | CXCL5 | -0.001 | 0.993 |
| Cytokines (pg/ml) | G-CSF | -0.164 | 0.236 |

### Table E6. Proteins in the leading edge Table

| | | |
|---|---|---|
| **REACTOME_CELLULAR_RESPONSES_TO_STIMULI** | HBB, HBA2, HSP90B1, GSR, CAT, TALDO1, PSMA1, PSMA5, TLN1, GPX3, PSMB6, BLVRB, PSMA4 | pre_vs_post |
| **REACTOME_CELLULAR_RESPONSE_TO_CHEMICAL_STRESS** | HBB, HBA2, GSR, CAT, TALDO1, PSMA1, PSMA5, GPX3, PSMB6, BLVRB, PSMA4 | pre_vs_post |
| **REACTOME_COMPLEMENT_CASCADE** | CFP, C4BPA, C4BPB, CFHR5, C1R, CFH, F2, PROS1, CFI, C6, CFHR2, C7, CFHR4, C3, CFHR1, CRP, MASP1, C8G, C1S, C8A | pre_vs_post |
| **REACTOME_FORMATION_OF_FIBRIN_CLOT_CLOTTING_CASCADE** | F13B, F10, KNG1, F2, KLKB1, PROS1, PRCP, F13A1, F5, F9, PROC, F7, F11, PROCR | pre_vs_post |
| **REACTOME_HEMOSTASIS** | F13B, F10, KNG1, F2, KLKB1, PROS1, APOH, PRCP, F13A1, F5, PLG, F9, A1BG, PROC, F7, ITIH4, SELL, YWHAZ, F11, IGF2, TTN, PROCR | pre_vs_post |
| **REACTOME_INNATE_IMMUNE_SYSTEM** | S100A9, LCN2, CFP, S100A7, CRISP3, S100A8, C4BPA, C4BPB, PGLYRP2, CFHR5, C1R, CFH, F2, PROS1, CFI, PRCP, C6, A1BG, LYZ, LRG1, CFHR2, C7, SAA1, FABP5, CFHR4, C3, CFHR1, JUP, CRP, SELL, MASP1, PRG2, CD14, C8G, C1S, DSP, C8A, CST3, CAMP, SERPINA3 | pre_vs_post |
| **REACTOME_INTRINSIC_PATHWAY_OF_FIBRIN_CLOT_FORMATION** | F10, KNG1, F2, KLKB1, PROS1, PRCP, F9, PROC, F11 | pre_vs_post |
| **REACTOME_FORMATION_OF_THE_CORNIFIED_ENVELOPE** | JUP, DSP, KRT5, KRT14, KRT1, KRT71, KRT3, KRT6A, KRT6B, DSG1, KRT10, KRT9, KRT2, KRT13, DSC2 | pre_vs_healthy |
| **REACTOME_INNATE_IMMUNE_SYSTEM** | LRG1, LYZ, CRP, S100A8, CFI, PRCP, S100A9, SAA1, SERPINA3, JUP, A1BG, DSP, C3, CFHR5, S100A7, RPS27A, HBB, CALML5, CAMP, CFB, GGH, CFHR3, B2M, CFHR1, KRT1, FCN3, HRNR, CST3, RNASE2, TXN, CPB2, CFH, DSG1, FABP5, CTSD, CD14, CD44, LBP, CRISP3, ITLN1, C4BPA, CFHR4, SERPING1 | pre_vs_healthy |
| **REACTOME_KERATINIZATION** | JUP, DSP, KRT5, KRT14, KRT1, KRT71, KRT3, KRT6A, KRT6B, DSG1, KRT10, KRT9, KRT2, KRT13, DSC2 | pre_vs_healthy |
| **REACTOME_NEUTROPHIL_DEGRANULATION** | LRG1, LYZ, S100A8, PRCP, S100A9, SERPINA3, JUP, A1BG, DSP, C3, S100A7, HBB, CALML5, CAMP, GGH, B2M, KRT1, HRNR, CST3, RNASE2, DSG1, FABP5, CTSD, CD14, CD44, CRISP3 | pre_vs_healthy |

**Table E7. Relationship of systemic neutrophilic inflammation with lung function**

| Dataset | Variable | Spearman Correlation | p value |
|---|---|---|---|
| Immune cells | Neutrophils | 0.041 | 0.775 |
| Neutrophil cell surface markers | CD101 | 0.056 | 0.713 |
| Neutrophil cell surface markers | CD45 | -0.270 | 0.058 |
| Neutrophil cell surface markers | CXCR4* | -0.444 | 0.001 |
| Neutrophil cell surface markers | HLA-DR* | -0.432 | 0.002 |
| Neutrophil cell surface markers | CD206 | -0.162 | 0.260 |
| Neutrophil cell surface markers | CD15 | -0.156 | 0.278 |
| Neutrophil cell surface markers | CD16* | -0.311 | 0.028 |
| Neutrophil cell surface markers | CCR7* | -0.349 | 0.013 |
| Neutrophil cell surface markers | CD66b | -0.272 | 0.056 |
| Cytokines | IL3 | 0.026 | 0.860 |
| Cytokines | IL6 | -0.144 | 0.322 |
| Cytokines | IL8 | -0.219 | 0.131 |
| Cytokines | G-CSF | -0.132 | 0.368 |
| Cytokines | CXCL5 | 0.252 | 0.081 |
| Cytokines | CXCL1 | 0.009 | 0.951 |

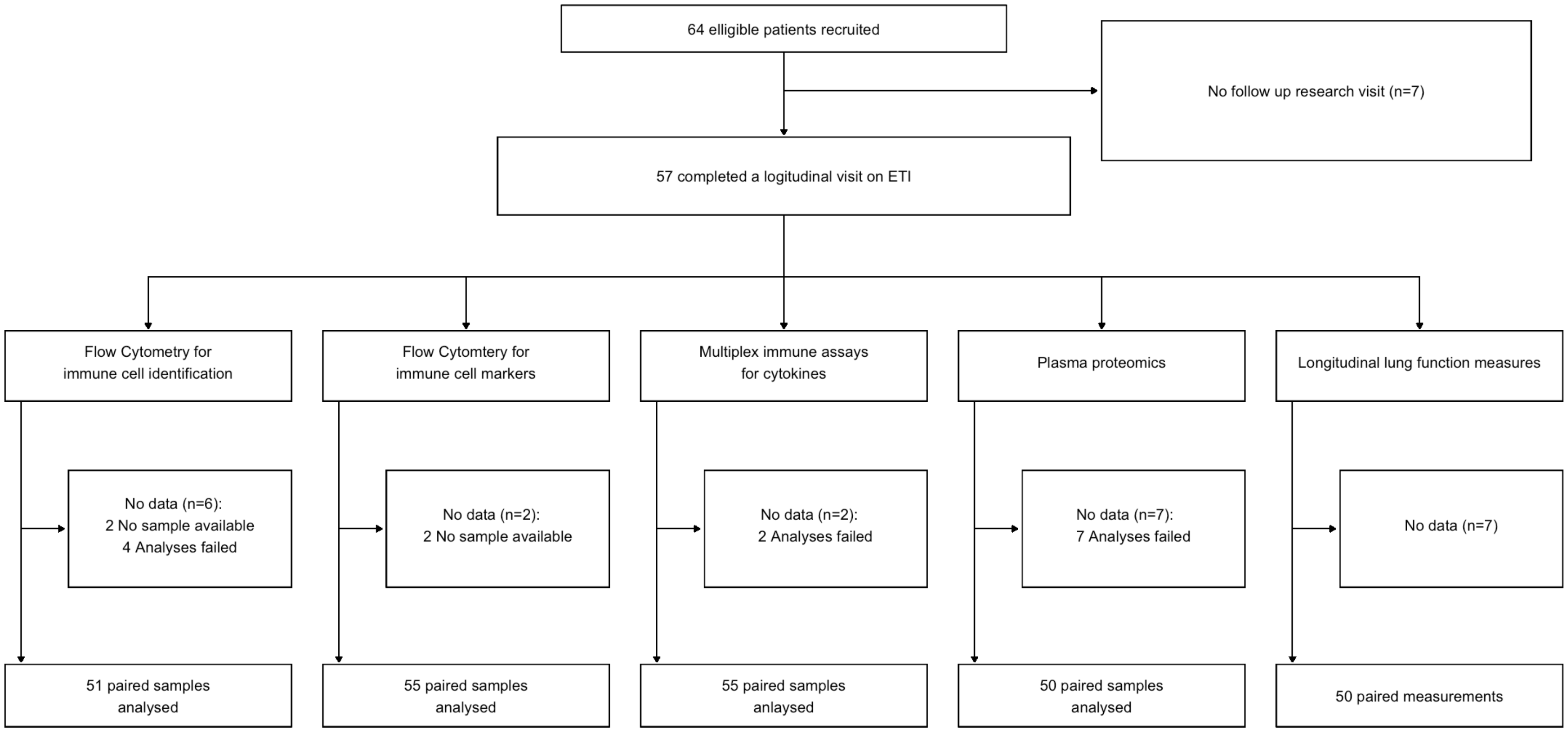


**Figure E1. Consort diagram for recruited patients**

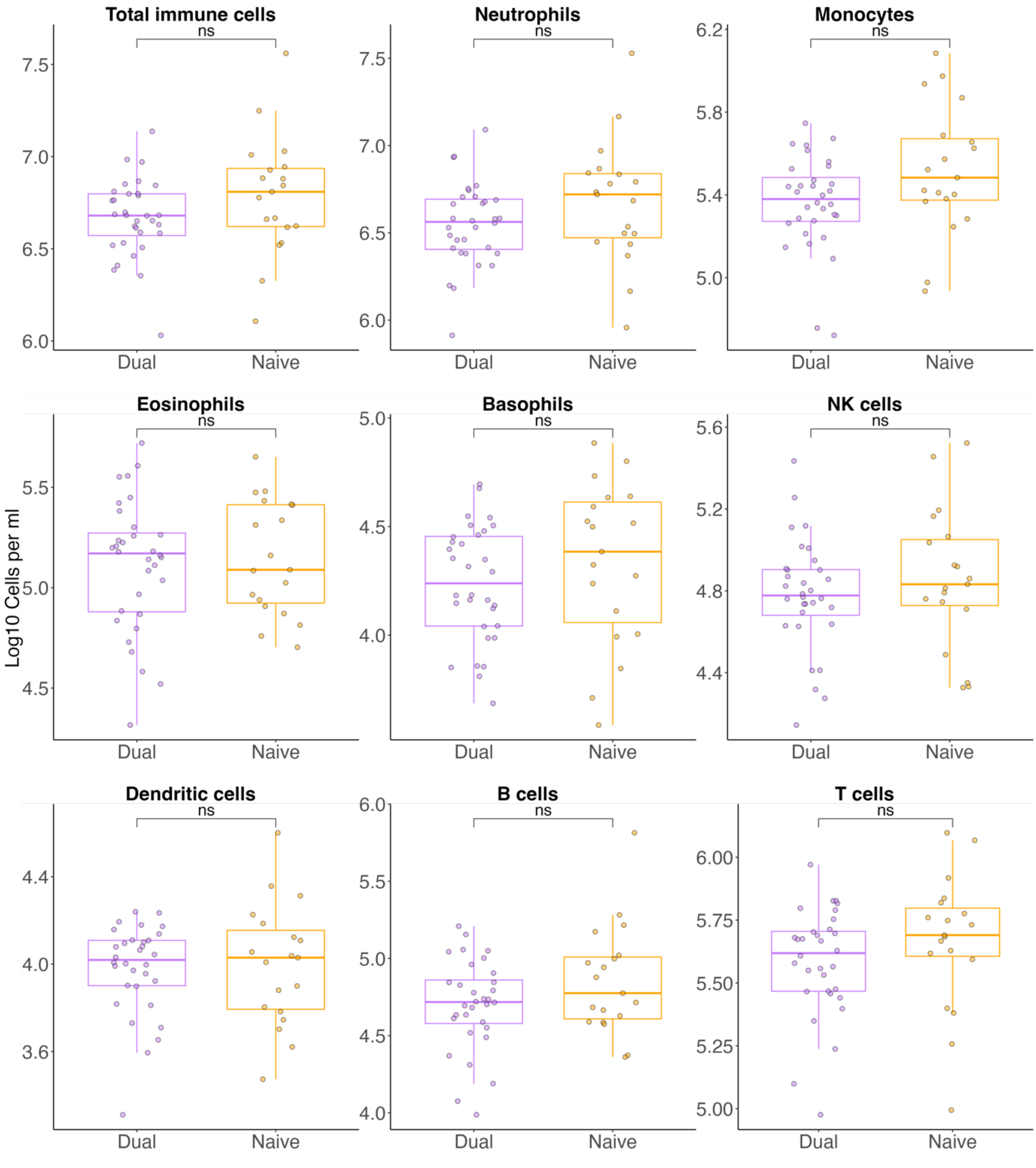


**Figure E2. Immune profiles of pre-ETI CF subjects stratified by baseline modulator status.** Absolute cell counts were measured by high-dimensional flow cytometry, with CF subjects stratified according to baseline modulator status (dual therapy vs modulator naïve). For each boxplot, individual cell count, together with median and interquartile ranges are plotted. Difference between cohorts are calculated by t-tests: **** = p < 0.0001, *** = p < 0.001, ** = p < 0.01, * = p < 0.05.

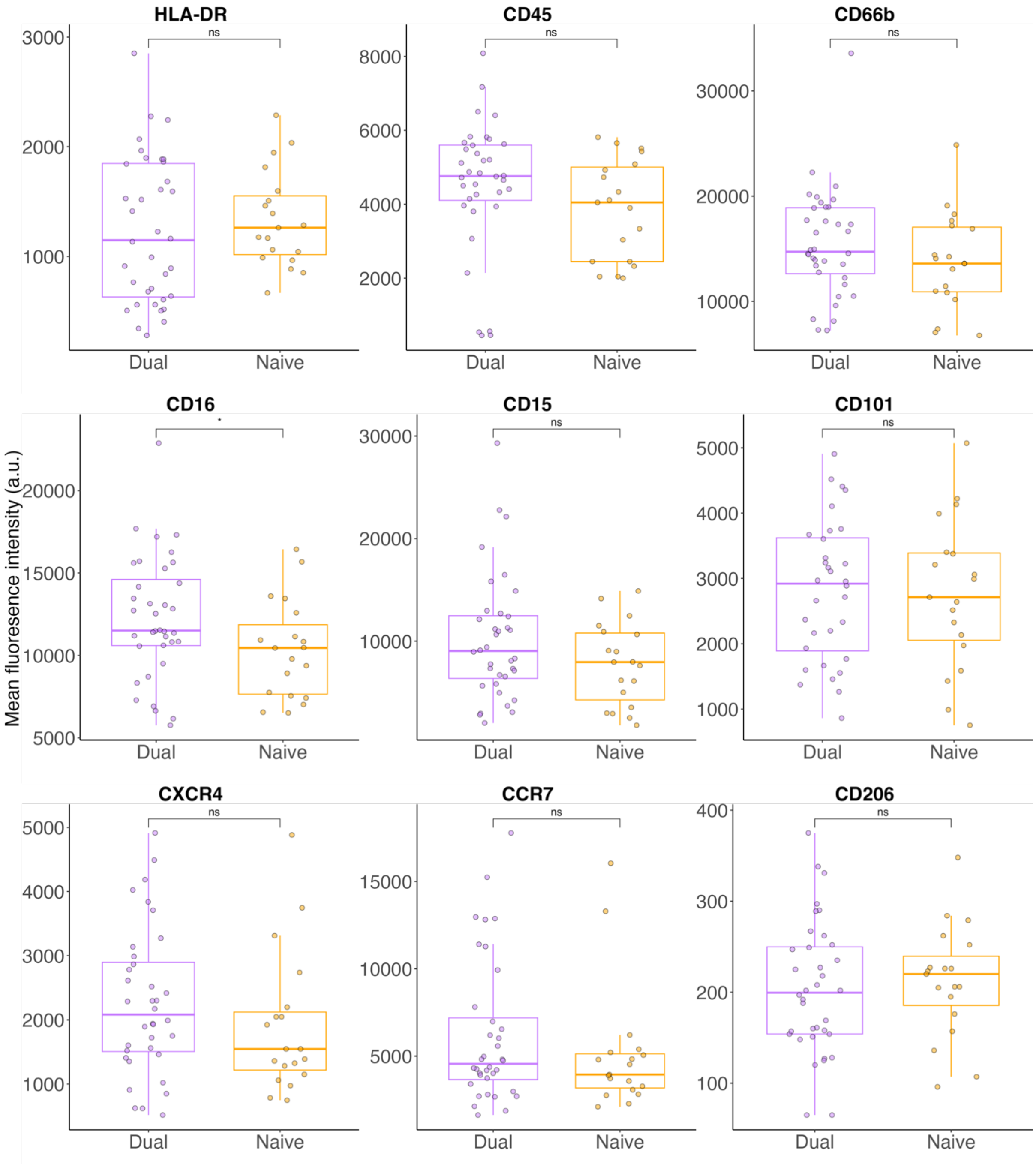


**Figure E3. Immunophenotyping of neutrophils of pre-ETI subjects stratified by baseline modulator status.** Median fluorescence intensity (MFI) was measured by high-dimensional flow cytometry. MFI values are plotted, with CF subjects stratified according to baseline modulator status (dual therapy vs modulator naïve). For each group, individual median cell marker MFI, together with median and interquartile ranges are plotted. Difference between cohorts are calculated by Mann Whitney: **** = $p < 0.0001$, *** = $p < 0.001$, ** = $p < 0.01$, * = $p < 0.05$.

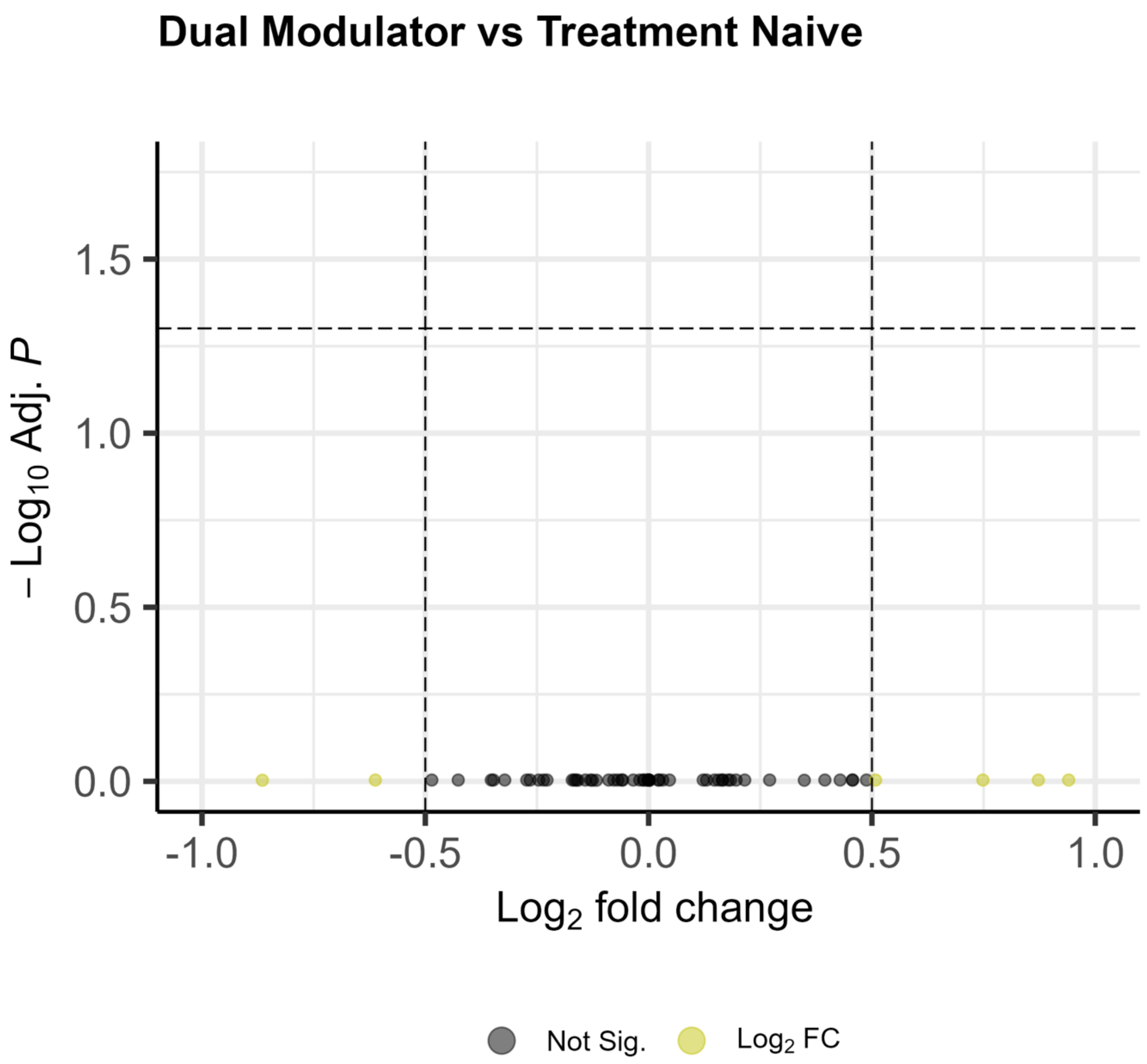


**Figure E4. Cytokines stratified by baseline modulator status.** Volcano plot for dual modulator versus treatment naive patients displaying fold change against adjusted p-values calculated by Mann Whitney. Dots are coloured by thresholds based on log-fold change and adjusted p value. Horizontal dashed line indicates cut off for adjusted p value < 0.05. Vertical dashed lined indicates positive and negative cut offs for absolute log2 fold change of 1.

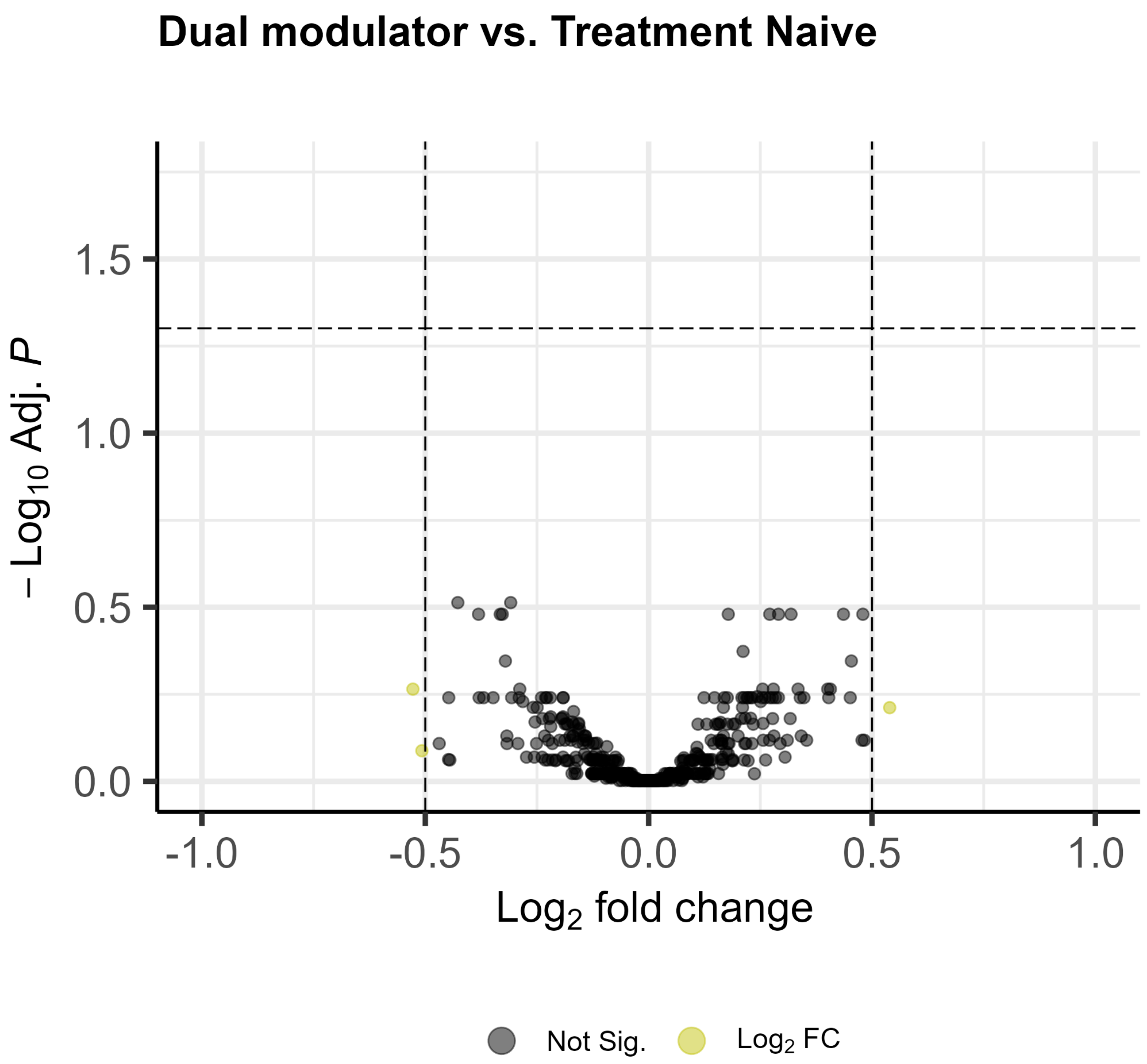


**Figure E5. Proteome stratified by modulator status.** Volcano plot for dual modulator versus treatment naive patients displaying fold change as calculated by limma against adjusted p-values. Dots are coloured by thresholds based on log-fold change and adjusted p value. Horizontal dashed line indicates cut off for adjusted p value < 0.05. Vertical dashed lined indicates positive and negative cut offs for absolute log2 fold change of 1.

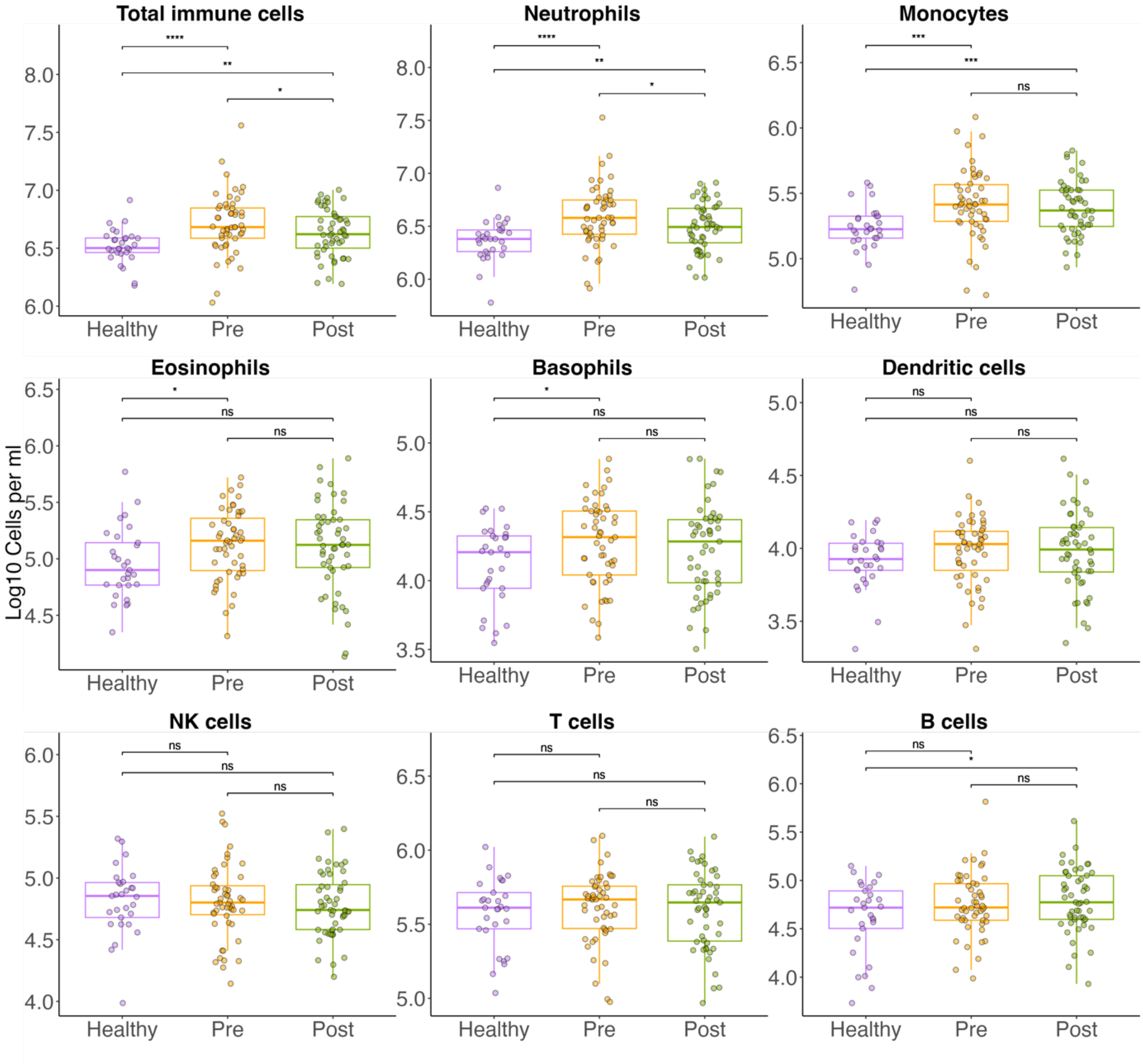


**Supplementary E6. Effect on ETI on Immune cell populations.** Absolute cell counts were measured by high-dimensional flow cytometry. (A) Stacked bar chart displaying median values of all circulating immune cells . (B) Box plots of the selected immune cells frequently implicated in CF lung disease pathology, with CF subjects stratified according to ETI status. (C) Box plots of the monocyte subsets. For each boxplot, individual cell count, together with median and interquartile ranges are plotted. Difference between cohorts are calculated by t-tests: **** = $p < 0.0001$, *** = $p < 0.001$, ** = $p < 0.01$, * = $p < 0.05$

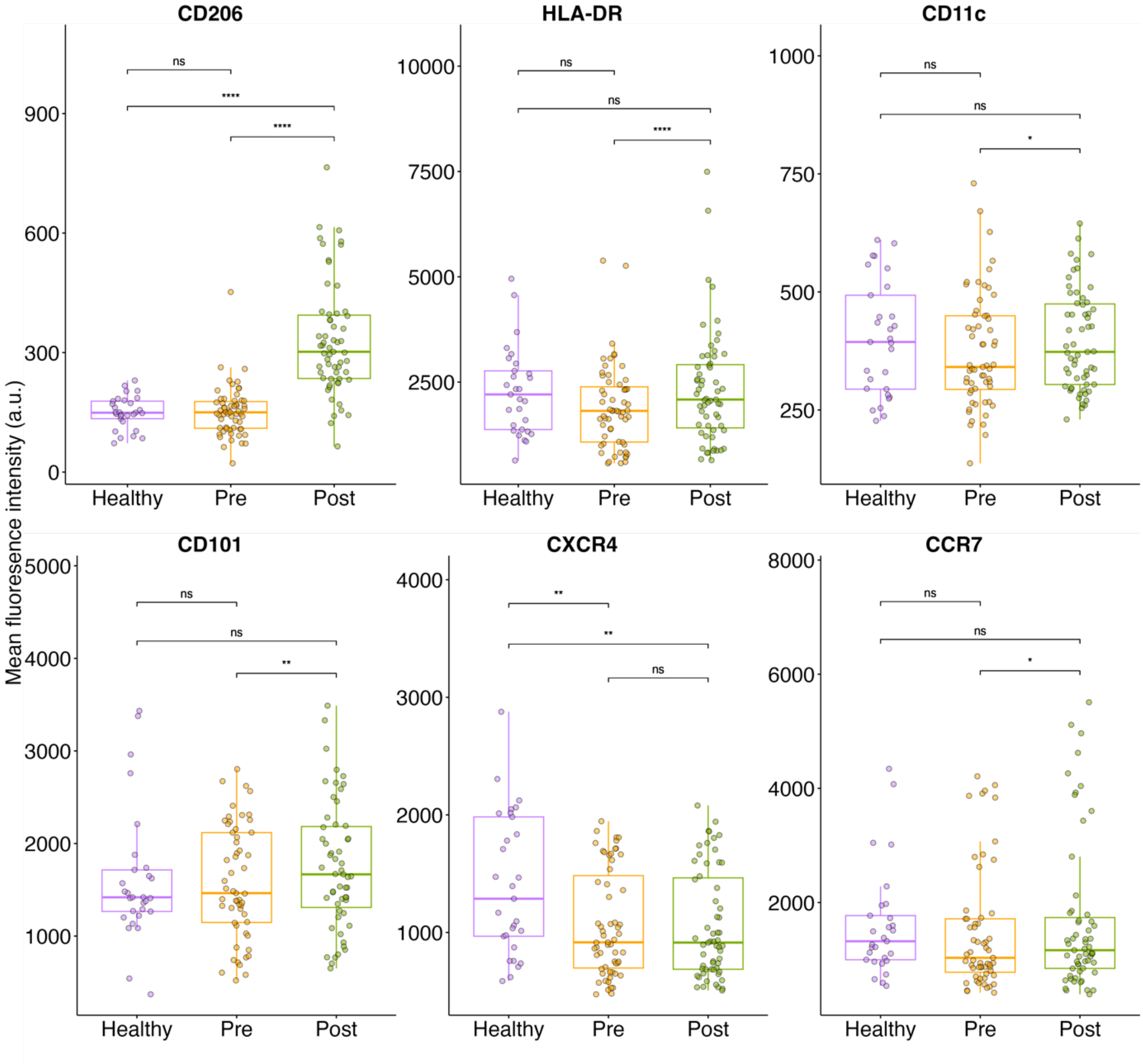


**Figure E7. Immunophenotyping of classical CD14+ monocytes pre- and post- ETI.** Median fluorescence intensity (MFI) was measured by high-dimensional flow cytometry. MFI values are plotted, with CF subjects stratified according to ETI status. For each group, individual median cell marker MFI, together with median and interquartile ranges are plotted. Difference between cohorts are calculated by Mann Whitney: **** = p < 0.0001, *** = p < 0.001, ** = p < 0.01, * = p < 0.05.

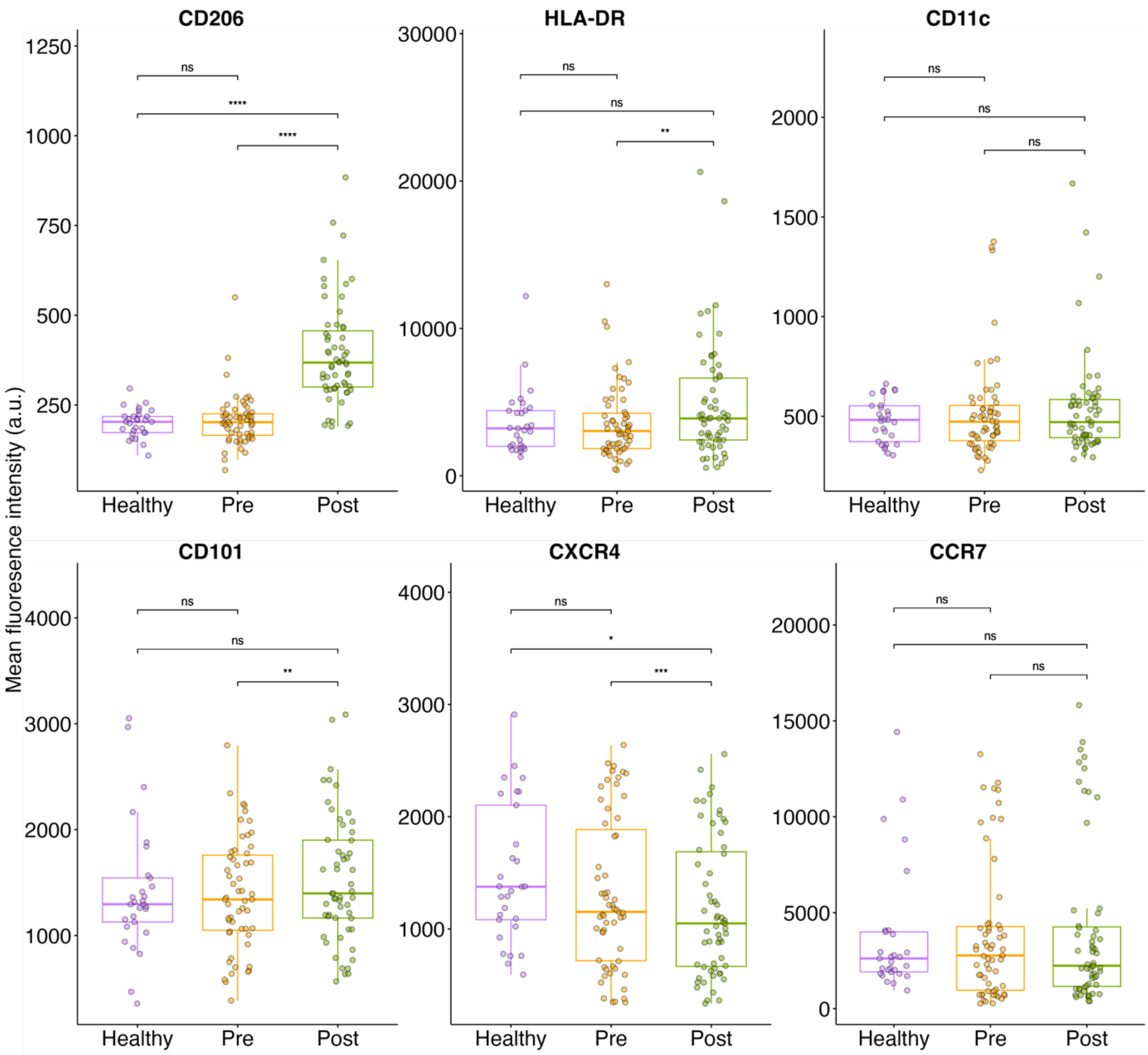


**Figure E8. Immunophenotyping of intermediate CD14+CD16+ monocytes pre- and post- ETI.** Median fluorescence intensity (MFI) was measured by high-dimensional flow cytometry. MFI values are plotted, with CF subjects stratified according to ETI status. For each group, individual median cell marker MFI, together with median and interquartile ranges are plotted. Difference between cohorts are calculated by Mann Whitney: **** = $p < 0.0001$, *** = $p < 0.001$, ** = $p < 0.01$, * = $p < 0.05$.

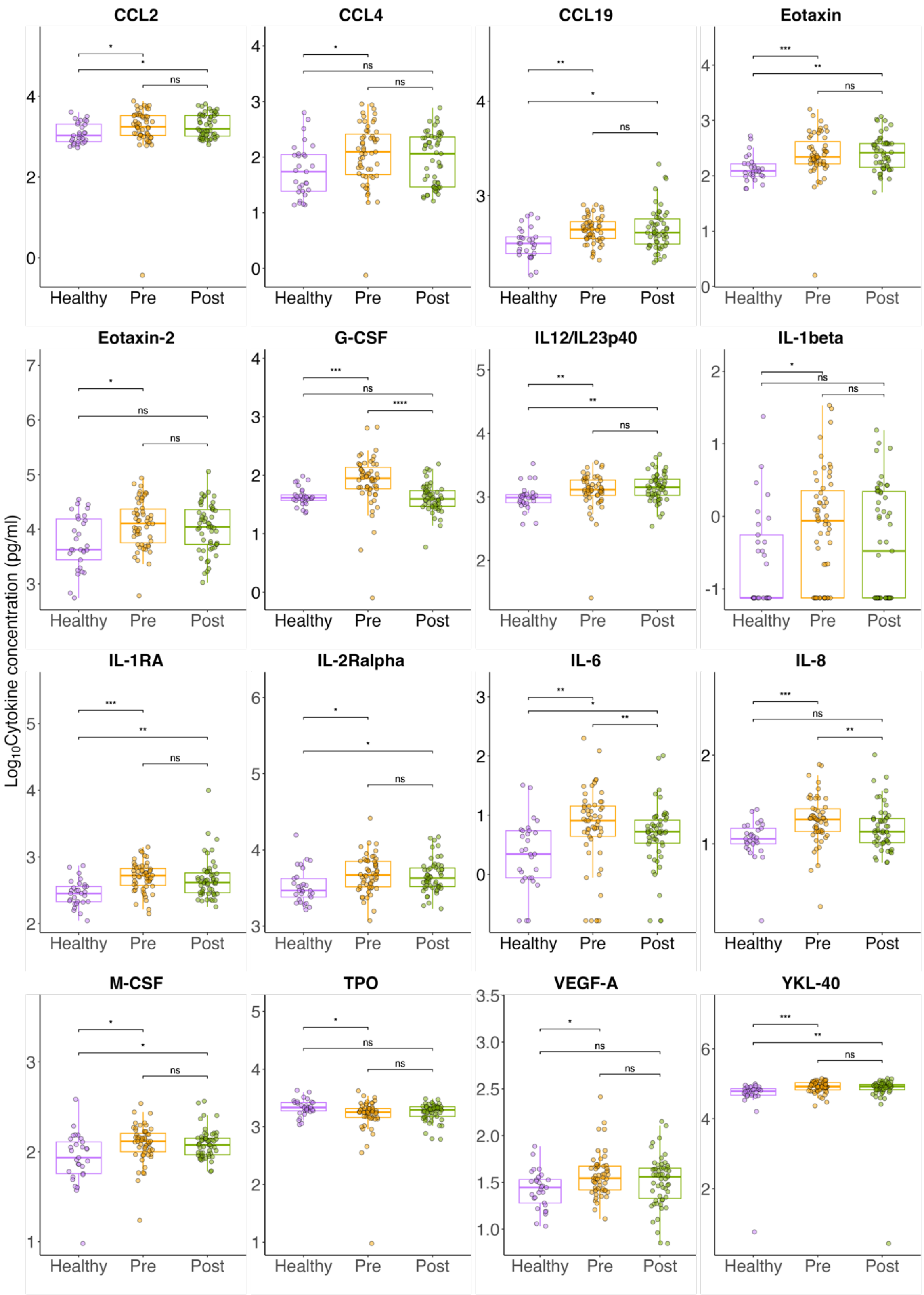


**Figure E9. Quantitive profiling of cytokines and chemokines.** For all those that were different at baseline between healthy and pre-ETI (adjusted p-value <0.05) the absolute concentrations are shown, with CF subjects stratified by ETI state. For each group, individual median cell marker MFI, together with median and interquartile ranges are plotted. Difference between cohorts are calculated by Mann Whitney: **** = $p < 0.0001$, *** = $p < 0.001$, ** = $p < 0.01$, * = $p < 0.05$.

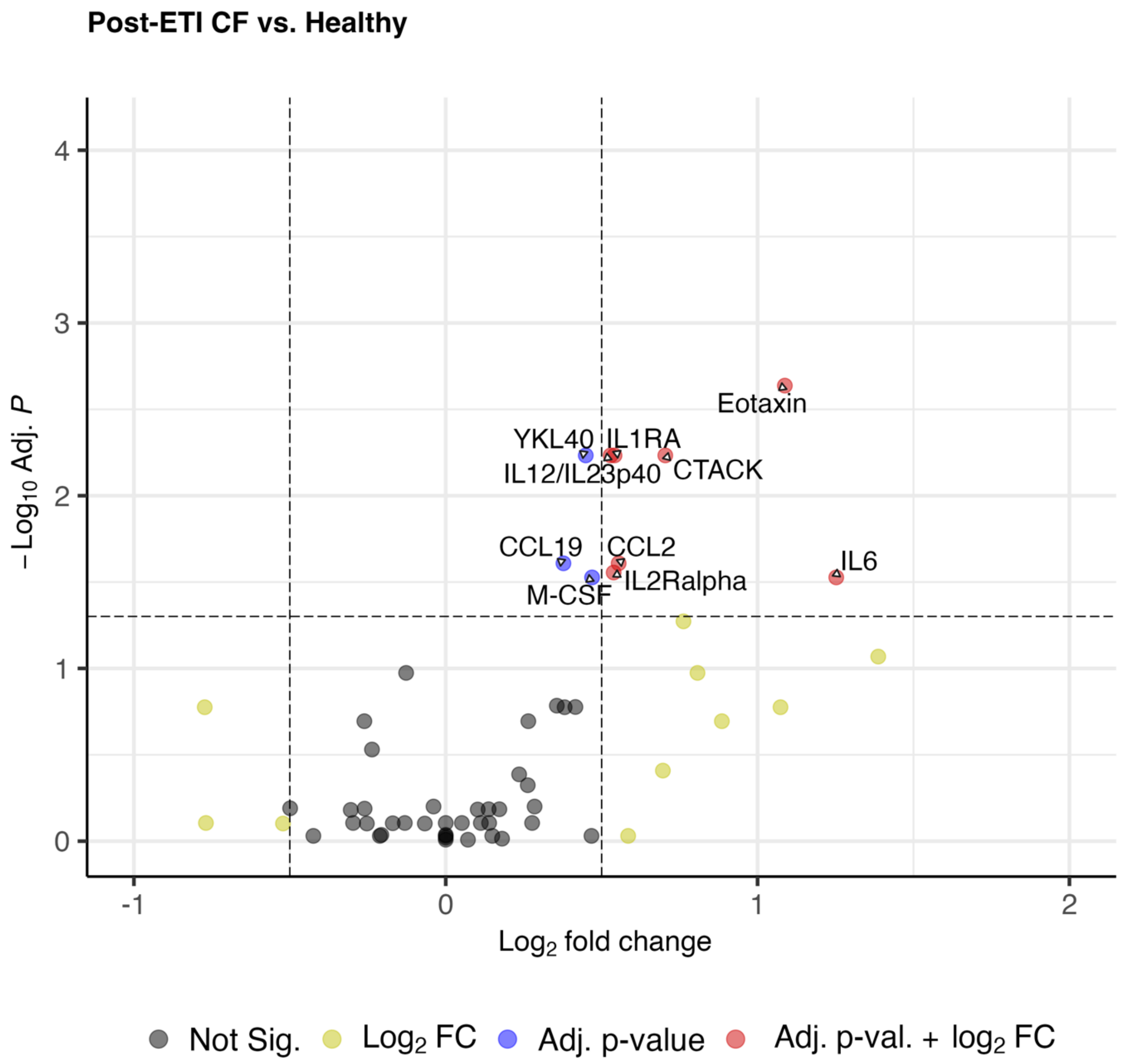


**Supplementary Figure E10. Quantative profiling of post-ETI systemic cytokines and chemokines.** Volcano plot displaying post-ETI CF versus healthy controls with log2median fold change in cytokine abundance against adjusted p-value calculated by Mann-Whitney U tests. Dots are coloured by thresholds based on log-fold change and adjusted p value. Horizontal dashed line indicates cut off for adjusted p value < 0.05. Vertical dashed lined indicates positive and negative cut offs for absolute log2 fold change of 1.

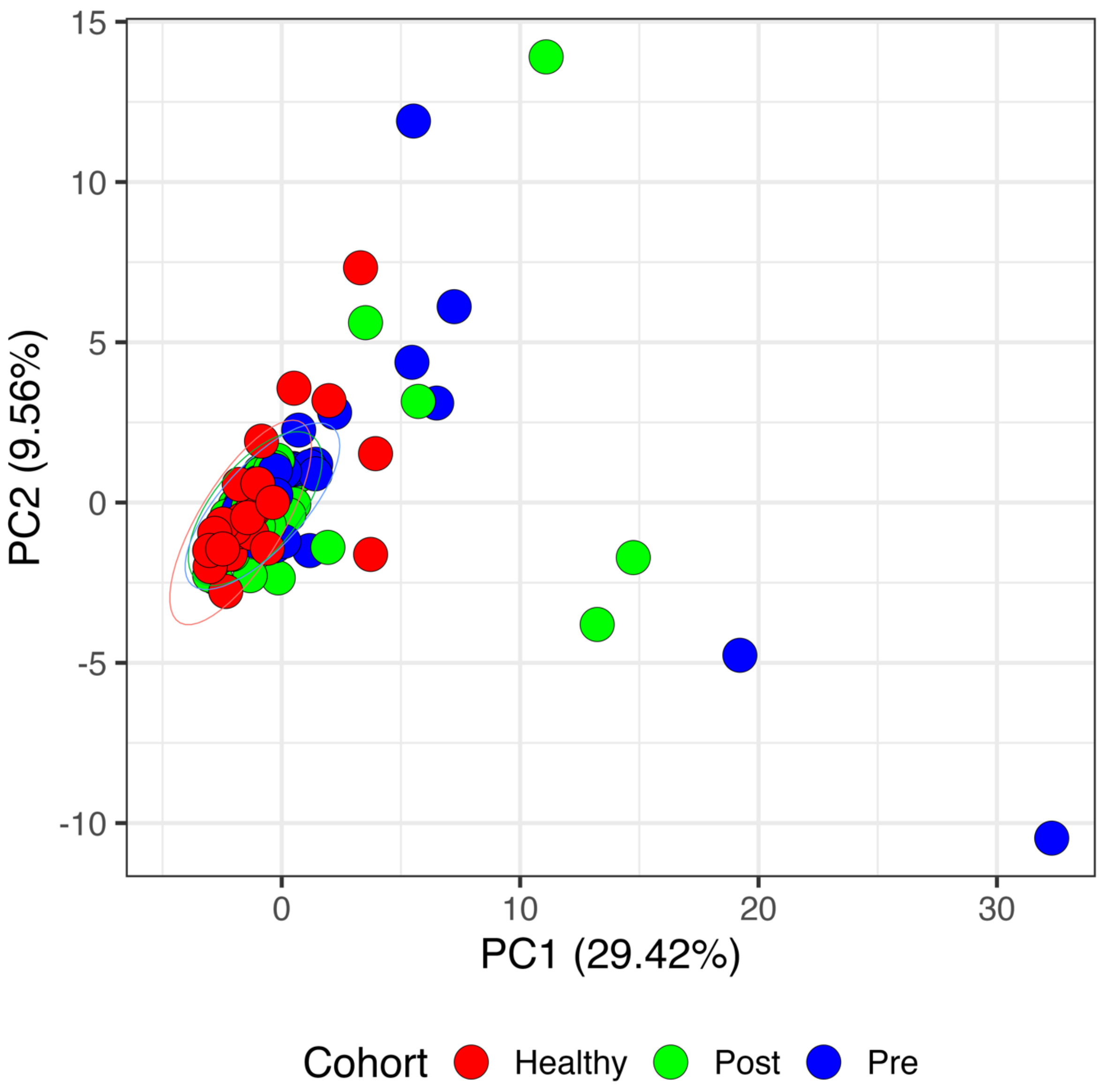


**Supplementary Figure E11. Principal component analysis of plasma cytokine and chemokine profiles.** The entire data set (55 CF subjects (110 paired samples) ; 29 healthy controls (29 samples)) were analysed by multiplex immunoassay (61 proteins) and subjected to principal component analysis (PCA). Samples were colour-coded by clinical cohort.

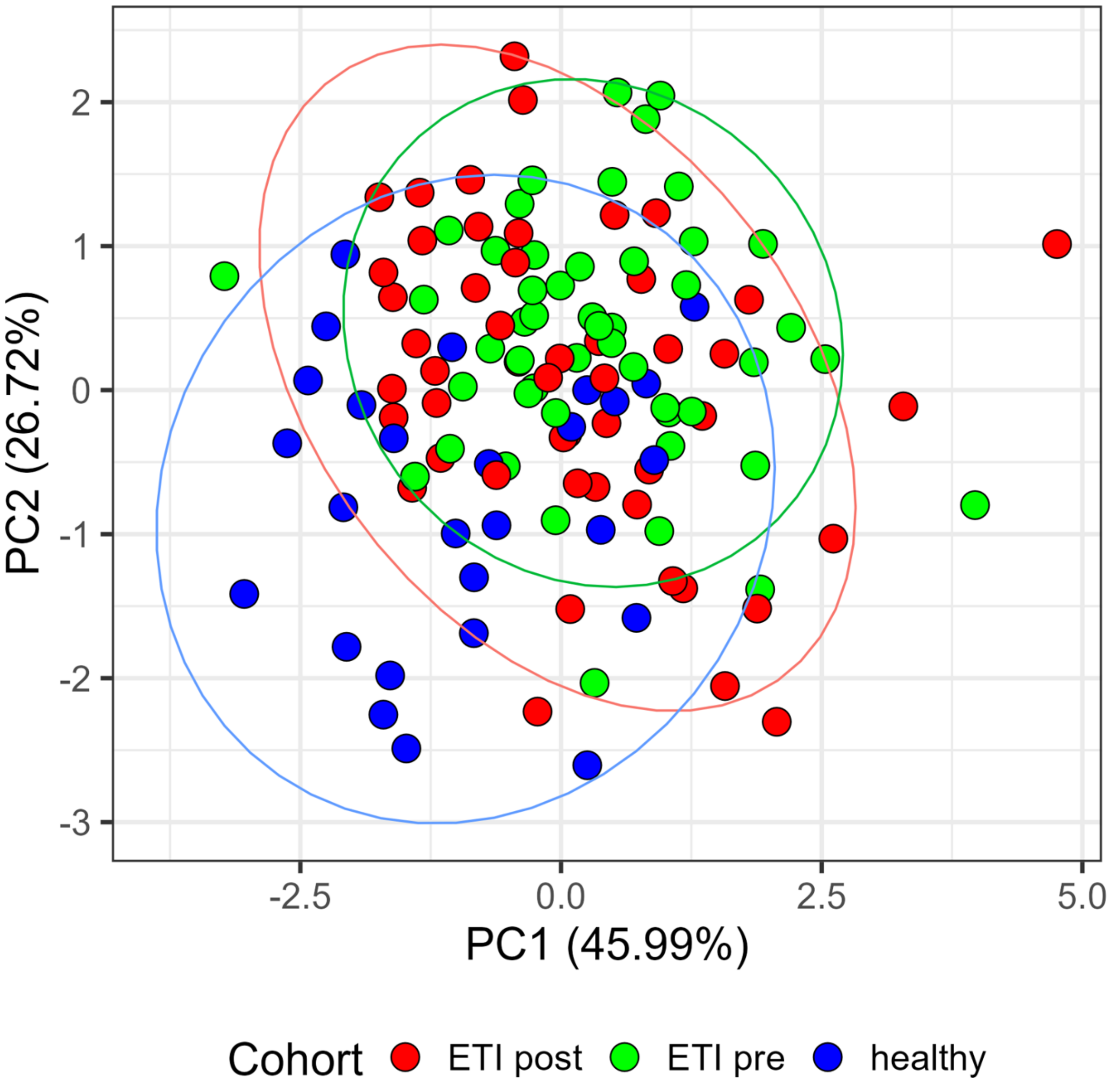


**Supplementary Figure E12. Principal component analysis of plasma protein profiles.** The entire data set (50 CF subjects (100 paired samples) ; 27 healthy controls (27 samples)) were analysed by tandem mass spectrometry and subjected to principal component analysis. Samples were colour-coded by clinical cohort.
